# Access Consequences of Restricting Mohs Micrographic Surgery to Fellowship-Trained Surgeons: A National Workforce, Board-Certification, and Drive-Time Simulation Study

**DOI:** 10.64898/2026.09.12.26362922

**Authors:** Henry Jeon, Mary E. Logue, George Murakawa, Kenneth K. Yu

## Abstract

**Background:** Policies restricting Mohs micrographic surgery (MMS) to only fellowship-trained (FT) surgeons, whether through privileging standards, laboratory-director qualifications, or payer credentialing, would bar residency/practice-trained (R/P-trained) surgeons from MMS. The access consequences have not been quantified.

**Objective:** To quantify the access consequences of restricting MMS to only FT surgeons.

**Methods:** Cross-sectional analysis of all Medicare MMS billers (2020-2024), classified FT or R/P-trained by audited American College of Mohs Surgery directory linkage and linked to Micrographic Dermatologic Surgery (MDS) certification; drive-time simulation of R/P-trained exclusion against the Centers for Medicare & Medicaid Services (CMS) dermatology access standard; fellowship-pipeline replacement modeling; and projection of rural Mohs capacity needed through 2041.

**Results:** R/P-trained surgeons were 45.3% of the 2024 workforce (55.6% MDS-certified), supplied 52.1% of nonmetropolitan MMS volume, and were the only Mohs surgeons in 223 counties. On the specialty’s quality measure, stages per case, the pathways were indistinguishable (median 1.5 in both). Excluding R/P-trained surgeons would put 23.1 million people newly outside the CMS dermatology access standard, with nonmetropolitan drive times rising by 3.2 times the metropolitan increase. R/P-trained surgeons perform 353,736 Medicare cases a year, and the remaining surgeons could not care for all of the displaced patients even at 50% higher caseloads. With R/P-trained surgeons grandfathered, 114 counties would still lose in-county access by 2041. At present, rural access falls 323 surgeons short of the CMS standard. Closing that shortfall by 2041 would take about 77 new rural Mohs surgeons a year, four times today’s entry rate.

**Limitations:** Fee-for-service claims; directory-based classification, audited and adjusted; no clinical outcomes.

**Conclusion:** A fellowship-only restriction would produce large access losses, geographically concentrated and disproportionately rural, and the fellowship pipeline could not replace the excluded surgeons in the counties that would lose them. The workforce loss would affect both metropolitan and nonmetropolitan patients, but with far more severe consequences for nonmetropolitan residents. Access to Mohs surgery rests on both training pathways. Policy should expand every route into Mohs surgery, through more fellowship positions, stronger Mohs training within dermatology residency, and a reopened practice pathway to MDS certification, and should keep laboratory-director and network eligibility keyed to the primary dermatology board certification common to both pathways.

**CAPSULE SUMMARY:**

- Privileging, CLIA laboratory-directorship, and payer credentialing policies could restrict Mohs surgery to only fellowship-trained surgeons; the workforce literature had not quantified the access consequences.
- Excluding residency/practice-trained surgeons would disproportionately affect nonmetropolitan patients, leave 223 counties without in-county Mohs access, push 23 million people outside the Centers for Medicare & Medicaid Services’ dermatology access standards, and displace roughly 212,000 cases per year that the remaining workforce could not absorb, lengthening waits in urban and rural areas alike; access policy should treat the two pathways as coequal routes into Mohs surgery, as Medicare coverage policy already does.

## INTRODUCTION

Skin cancer is the most common cancer in the United States, with more than five million keratinocyte carcinomas treated in over three million people each year, and incidence continues to rise.^1^ For high-risk tumors, and for tumors of the face, ears, hands, and other sites where tissue conservation and margin control matter most, the treatment of choice is Mohs micrographic surgery (MMS).^2–4^ MMS is performed by dermatologists who enter the field through fellowship training (American College of Mohs Surgery [ACMS]-affiliated and, since 2003, Accreditation Council for Graduate Medical Education [ACGME] Micrographic Surgery and Dermatologic Oncology [MSDO] fellowships) or through residency-based and structured practice-based training (courses, preceptorships, and proctored experience, hereafter residency/practice-trained [R/P-trained]).^5–7^ Both groups bill the same procedure codes and treat the same cancers. No comparative clinical-outcome study of the two groups exists, and on the specialty’s standard quality measure they are indistinguishable (within 0.05 stages per case).^8–10^ Because the Mohs surgeon acts as both surgeon and pathologist, with an on-site laboratory federally regulated as high-complexity histopathology,^11–13^ the ability to practice MMS is unusually dependent on credentialing determinations, namely privileges, laboratory directorship, and payment for the procedure codes.

Several instruments could restrict MMS to only fellowship-trained surgeons. The 2023 Clinical Laboratory Improvement Amendments (CLIA) personnel rule inadvertently removed the laboratory-director qualification route most dermatologists used. The Centers for Medicare & Medicaid Services (CMS) restored eligibility on pathway-neutral, certification-based terms,^12–15^ and a 2026 CMS/CDC request for information notes inquiries about adding “Mohs testing” as a CLIA specialty, a potential vehicle for training-based, fellowship-gated personnel standards.^16^ Payer network credentialing, or payment rules tied to the Mohs procedure codes (Current Procedural Terminology [CPT] 17311-17315), could effect the same restriction contract by contract. Facility privileging can impose it one institution at a time, and the Veterans Health Administration (VHA) has imposed it. Since 2023 the VHA has required fellowship training of surgeons performing MMS in its facilities. An R/P-trained surgeon may start or continue performing it only by having passed the Micrographic Dermatologic Surgery (MDS) subspecialty examination by the end of 2023 and by an individually granted facility waiver.^17,18^ That rule ranks fellowship training above the subspecialty board certification itself, since the fellowship graduate needs nothing more and the MDS-certified R/P-trained surgeon still needs a waiver. It narrowed the pool of board-certified surgeons who may treat veterans in VHA facilities, without any demonstrated benefit. No comparative outcome study supported it then, and none supports it now. The first nationwide assessment of MMS within the VHA found no difference in stages per case by ACMS membership,^8^ and the rule’s cost to veterans’ access has not been reported. In whichever setting it applies, each of these instruments would have the same effect, that R/P-trained surgeons would largely stop performing MMS there. The consequences would reach beyond the surgeons themselves. Practices would close, and the careers that residents and young surgeons intended to build through residency-and practice-based training would be foreclosed. This study simulates the exclusion at national scale and measures the consequence that falls on patients, their access to care.

The published workforce literature describes a heavily metropolitan MMS workforce,^19–22^ characterizes surgeons by society membership and certification status,^9,10,23^ and documents fellowship graduates’ metropolitan practice locations.^24^ A communication published after this analysis was completed confirmed that most rural Mohs billers are not listed by the ACMS.^25^ The literature therefore leaves unanswered what restricting MMS to fellowship-trained surgeons would mean for access (Table V): (1) how much MMS care each training pathway delivers, nationally and by rurality; (2) which counties an exclusion would leave with no Mohs surgeon; (3) how much patient travel time would rise, and how the increase would fall on residents by their rurality; (4) whether the remaining surgeons could absorb the displaced cases; (5) whether the excluded surgeons hold subspecialty certification; and (6) whether the fellowship pipeline could replace both the excluded surgeons and the workforce lost each year to retirement and other attrition. We set out to answer those questions for the entire Medicare Mohs workforce. The census covers every surgeon who billed Medicare for MMS from 2020 through 2024, the drive-time simulation covers the national population, and the replacement model runs to 2041. The answers raise a further question, how much rural Mohs capacity will be needed to keep today’s access as the rural population ages, and the study closes by estimating that need.

## METHODS

Full methods, all parameters, documentation of artificial intelligence (AI)-assisted analytic execution (S-M9), and a Strengthening the Reporting of Observational Studies in Epidemiology (STROBE) checklist for cross-sectional studies^26^ appear in the Supplementary Material.

### Workforce census

The census counted as an active Mohs surgeon any individual National Provider Identifier (NPI) that billed the first-stage MMS codes (CPT 17311/17313) in the CMS Medicare Physician & Other Practitioners public use files (PUF) for 2020-2024, with the stage codes 17312/17314/17315 contributing volumes.^27^ Everyone who billed in any year formed a dynamics roster (n=3,451) for the attrition and pipeline analyses, and those who billed in 2023-2024 formed a simulation roster (n=3,167) for the access simulation. Every practice location was geocoded to its county and ZIP Code Tabulation Area (ZCTA). Because the PUF suppresses any clinician-code-year with fewer than 11 beneficiaries, an estimated 124 surgeons (about 4%), plausibly rural and R/P-trained, are invisible to the census, and their absence biases the access losses reported here downward.

### Training-pathway classification

Consistent with prior national analyses,^9,10,19^ we defined fellowship training as listing in the ACMS directory, captured completely by a 133-seed ZIP-radius harvest and a 1,728-query surname sweep with tiered matching and logged adjudications. Every other biller was classified R/P-trained, and the American Society for Mohs Surgery (ASMS) public locator supplied an affiliation flag only. An independent surname-method count of ACMS-listed 2024 billers differed from ours by one surgeon.^28^ Because directory listing is a membership proxy for training, we audited a seeded sample of 30 R/P-classified surgeons for residual fellowship-trained misclassification and re-derived every principal result under the resulting training-based adjustment (Table VI).

### Board certification

All 3,451 NPIs were checked against the American Board of Dermatology’s (ABD) verification service, supplemented by strictly matched public MDS diplomate lists (2021-2025; 3,013 unique names),^29,30^ with unresolved cases counted as uncertified, so the certification shares reported are minimums.

### Geography

Counties (2023 vintage) carried US Department of Agriculture (USDA) Rural-Urban Continuum Codes (RUCC; nonmetropolitan = 4-9),^31^ population, Medicare enrollment, median household income, and primary-care Health Professional Shortage Area (HPSA) status. County access classes (R/P-only, FT-only, mixed) were computed under the primary and adjusted classifications, and Hospital Service Area (HSA) and Hospital Referral Region (HRR) analyses tested boundary sensitivity.

### Exclusion simulation

Road-network drive times (Open Source Routing Machine [OSRM]) ran from 26,419 populated-ZCTA population-weighted centroids (331.4 million people; 68.1 million beneficiaries) to the nearest surviving Mohs practice site under the status quo (A), under exclusion of all 1,474 R/P-trained simulation-roster surgeons (B), and under exclusion with the adjusted classification (B2). These runs yielded drive-time distributions, the populations beyond 30, 60, 90, and 120 minutes, compliance with the dermatology time-and-distance standards CMS applies to Medicare Advantage networks (42 CFR 422.116),^32^ and the capacity-constrained absorption of the excluded volume (B3), in which each surviving FT surgeon’s caseload was assumed to expand by 20%, with 0% and 50% as sensitivity values.

### Replacement model

To ask whether the fellowship pipeline could replace excluded rural capacity, we built a rural-capacity cohort model from MSDO pipeline supply (ACGME, SF Match, ACMS reports),^33–35^ graduates’ nonmetropolitan location rates, and graduation-year-based ages, and applied it identically to no restriction, to immediate exclusion, and to a restriction with grandfathering, in which existing R/P surgeons stay but R/P entry closes. In the primary analysis surgeons leave practice at the exit hazards measured in the 2021-2023 panel, before and after age 65, and retirement at a fixed age (65; 62 and 68) is the sensitivity analysis. County-level loss of in-county access under grandfathering uses the same attrition assumptions and an incumbent-based replacement channel, and a threshold analysis asked what nonmetropolitan-location rate among graduates would restore the 2026 rural headcount within 5, 10, and 15 years.

### Need

The replacement model holds demand at its 2024 level and takes today’s rural access as the target. We estimate need through 2041 in the sense used in health workforce planning, as the rural Mohs capacity that would keep status quo access and bring rural residents inside the CMS network-adequacy standard. Status quo access means rural in-county Mohs surgery at today’s rate per beneficiary as rural beneficiaries age and Mohs use per beneficiary grows. That standard is the set of dermatology time-and-distance maxima CMS applies to Medicare Advantage networks, graded by county type from 20 minutes and 10 miles in large metropolitan counties to 110 minutes and 100 miles in the sparsest,^32^ derived by mapping beneficiaries against provider locations so that the maxima reflect prevailing patterns of care rather than a clinical threshold.^36^ No Mohs-specific standard exists. Need in a year is the sum of two components. The first is status quo access, the 2026 rural in-county volume of the replacement model (53,232 cases). The second is the access gap, the first-stage volume that nonmetropolitan beneficiaries living outside the standard would generate at national age-specific rates. Both components were scaled for the projected change in rural fee-for-service beneficiaries by age band, carrying CY2025 county enrollment^37^ forward with published county projections,^38^ and for growth in cases per beneficiary of 3.2% per year, the rate at which all skin-cancer procedures per fee-for-service beneficiary grew over 2006-2012,^1^ which we chose as a deliberately restrained rate. The faster 2013-2024 trends in Mohs cases per fee-for-service beneficiary (5.6% per year, or 4.4% with the fee-for-service share held constant),^39^ the all-beneficiary trend (1.4%), and aging alone appear as sensitivities (Supplementary Material S-M10). Need converts to surgeons at the 2024 mean rural caseload (197.9 cases), and the surgeon count is a lower bound because rural sites rarely reach that caseload. The entry required is the constant annual rural entry that brings projected volume up to need by 2041 in the replacement model. Further sensitivities reduced all case counts by 5% and 10%, substituted state projections,^40^ used fixed-age retirement, let caseload grow 2% a year, and counted in the gap only beneficiaries more than two hours’ drive from a Mohs surgeon. For the population outside the standard, a county’s practice sites drop out in the year it loses its last Mohs surgeon. Need so defined is capacity that would have to exist within reach, not clinical need, and the residents it counts may obtain care today by traveling farther than the standard allows.

## RESULTS

### Workforce and certification (Table I)

In 2024, 3,042 surgeons billed Medicare fee-for-service for more than 1.1 million first-stage MMS cases. Of these, 1,665 (54.7%) were classified FT and 1,377 (45.3%) R/P-trained. The R/P-trained group billed 31.3% of national first-stage volume, and their median annual caseload was smaller, 168 cases against 395. On the specialty’s standard quality measure, however, the two groups were indistinguishable. That measure is stages per case, the number of tissue layers removed and examined before a tumor’s margins are clear, adopted as the specialty’s marker of surgical judgment because excess stages mean additional surgery and cost for the patient,^41^ and the median was 1.5 in both pathways.

**Table I.**
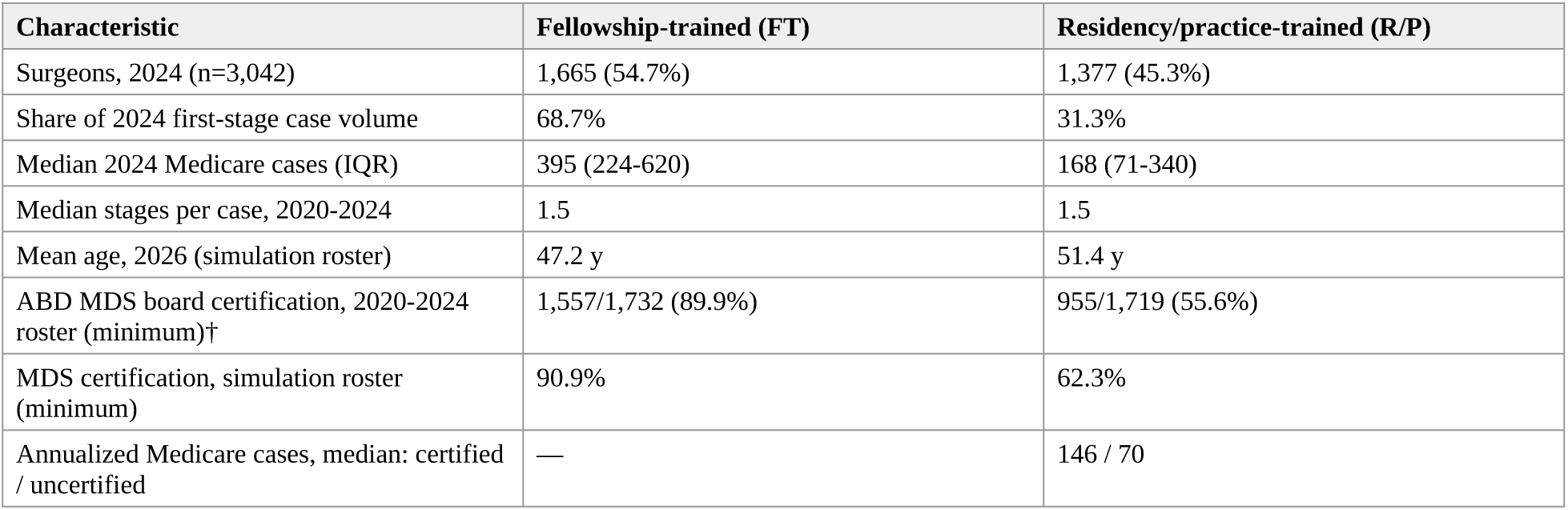
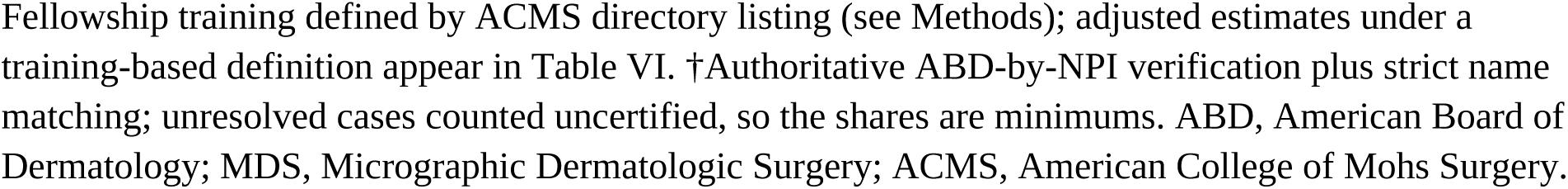
The US Medicare Mohs surgery workforce by training pathway, 2024 and 2020-2024.

| Characteristic | Fellowship-trained (FT) | Residency/practice-trained (R/P) |
| --- | --- | --- |
| Surgeons, 2024 (n=3,042) | 1,665 (54.7%) | 1,377 (45.3%) |
| Share of 2024 first-stage case volume | 68.7% | 31.3% |
| Median 2024 Medicare cases (IQR) | 395 (224-620) | 168 (71-340) |
| Median stages per case, 2020-2024 | 1.5 | 1.5 |
| Mean age, 2026 (simulation roster) | 47.2 y | 51.4 y |
| ABD MDS board certification, 2020-2024 roster (minimum)† | 1,557/1,732 (89.9%) | 955/1,719 (55.6%) |
| MDS certification, simulation roster (minimum) | 90.9% | 62.3% |
| Annualized Medicare cases, median: certified / uncertified | — | 146 / 70 |

Fellowship training defined by ACMS directory listing (see Methods); adjusted estimates under a training-based definition appear in Table VI. †Authoritative ABD-by-NPI verification plus strict name matching; unresolved cases counted uncertified, so the shares are minimums. ABD, American Board of Dermatology; MDS, Micrographic Dermatologic Surgery; ACMS, American College of Mohs Surgery.

Certification and fellowship are often conflated, but they describe different populations. Across the 2020-2024 roster, 89.9% of FT and 55.6% of R/P-trained surgeons held ABD MDS certification. A majority of the surgeons a fellowship-only rule would exclude are therefore subspecialty board-certified, and 38.0% of all certified billers are not ACMS members. R/P-trained diplomates outnumbered FT diplomates in the 2022, 2023, and 2025 certification cohorts.

### Who provides rural Mohs care (Table II; Figures 1-2)

The FT share of the 2023-2024 workforce fell steadily with rurality, from 58.4% in the largest metropolitan counties (RUCC 1) to 25.5% in RUCC 7. In the most rural counties (RUCC 9), all Mohs surgeons were R/P-trained. R/P-trained surgeons were the nonmetropolitan majority, 155 of 269, and performed 52.1% of 2024 nonmetropolitan volume. The odds of an R/P-trained surgeon practicing nonmetropolitan were 3.86 times those of an FT surgeon (95% CI 2.62-5.68; P<.001). An independent cross-section of 2023 billing that classified rurality by ZIP-code commuting area rather than county found the same composition, with 70.4% of rural Mohs billers not ACMS-listed against 44.1% of urban billers.^25^

**Figure 1.**
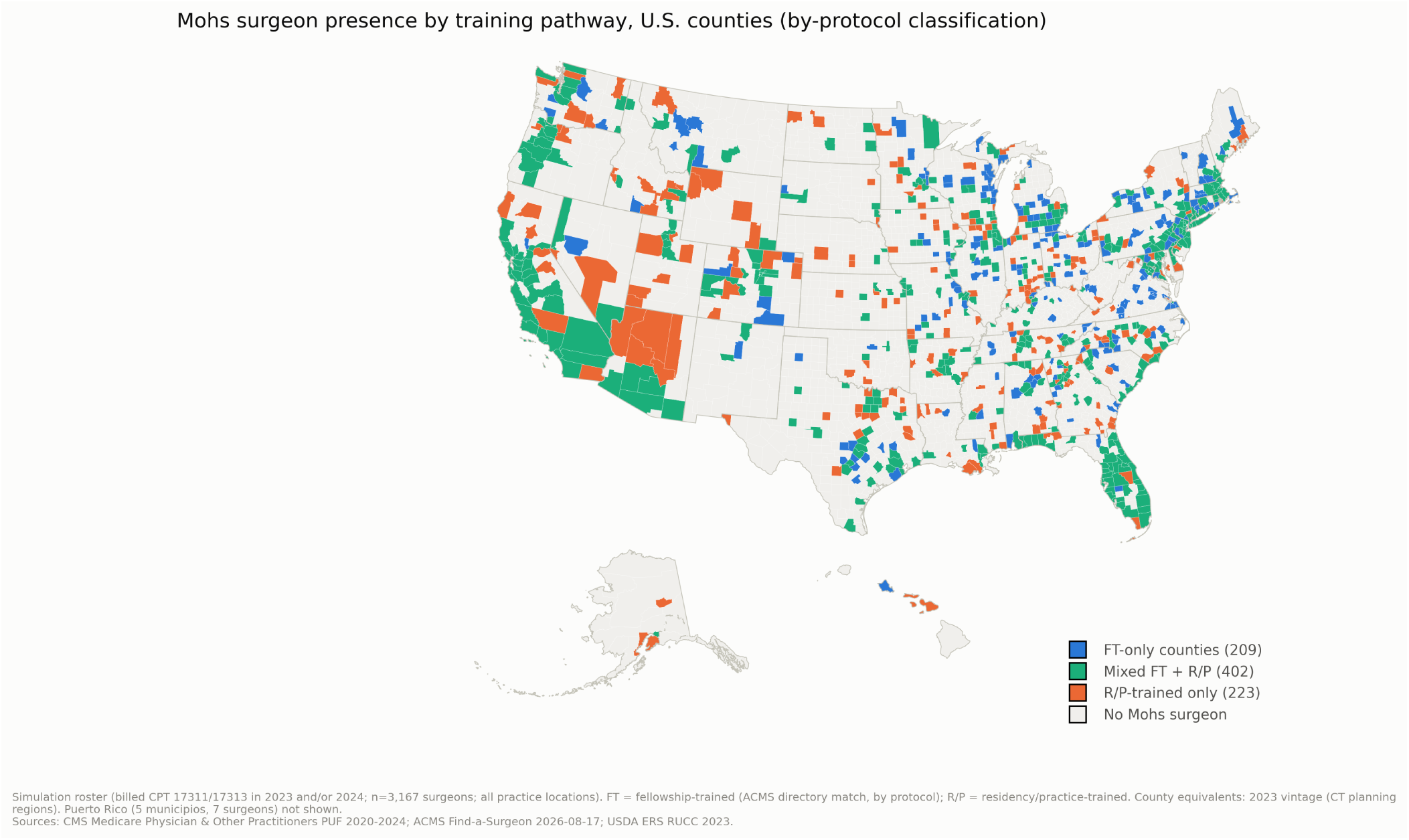
County-level Mohs surgery workforce by training pathway, 2023-2024. County access classes (fellowship-trained only, residency/practice-trained only, mixed, none) for the simulation roster (n=3,167), by-protocol classification. Adjusted-classification ranges in Table II.

**Figure 2.**
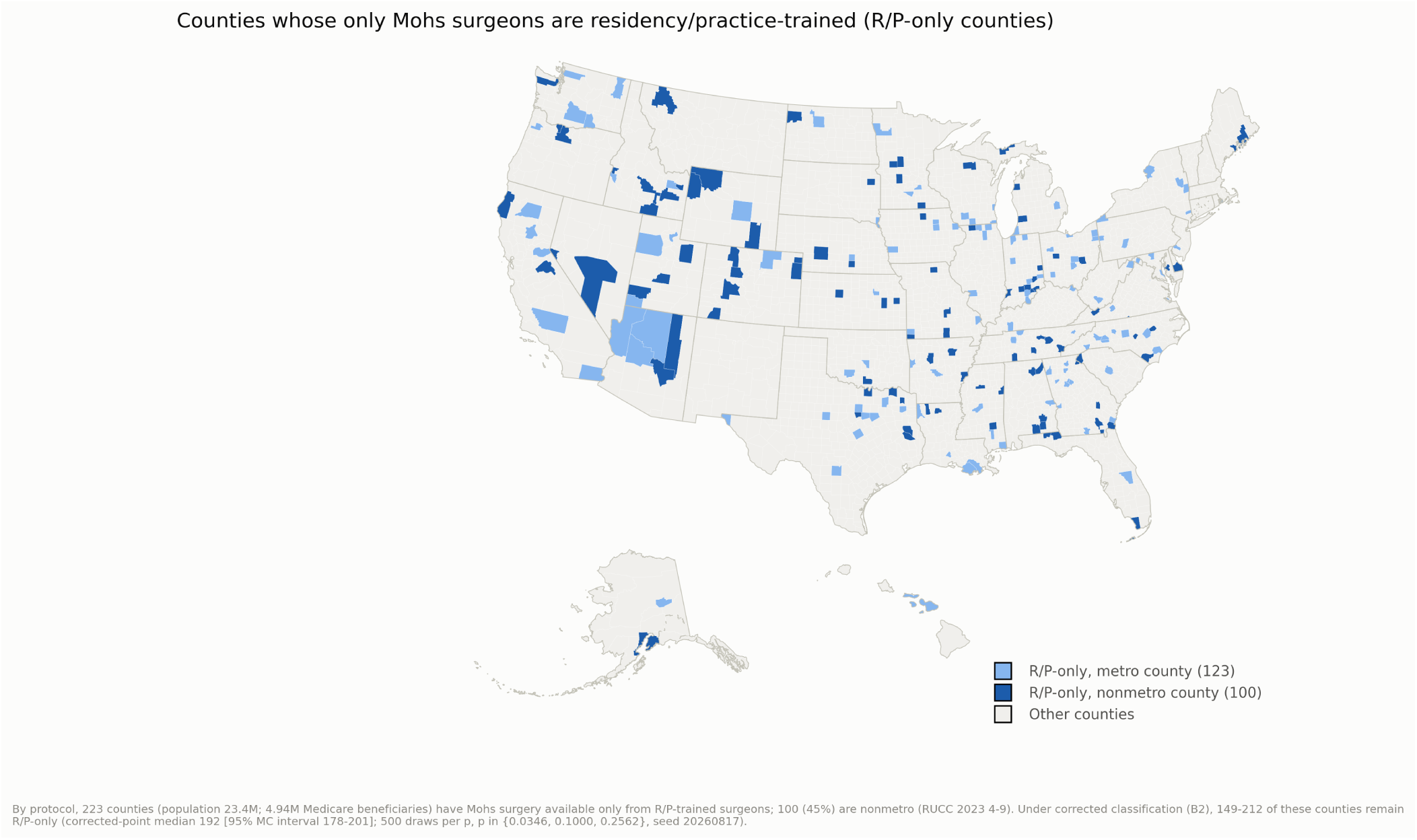
Counties whose entire Mohs workforce is residency/practice-trained. The 223 R/P-only counties (23.4 million residents), shaded by metropolitan/nonmetropolitan status. Under the training-based adjusted definition, 149-212 of these counties remain R/P-only (Table VI).

**Table II.** Geographic distribution and access classes (simulation roster, n=3,167)

| Measure | Value |
| --- | --- |
| FT share, metropolitan (RUCC 1-3) | 54.4% |
| FT share, nonmetropolitan (RUCC 4-9) | 42.4% |
| FT share by RUCC: 1 / 7 / 9 | 58.4% / 25.5% / 0 of 3 |
| R/P share of nonmetropolitan surgeons | 155/269 (57.6%) |
| R/P share of 2024 nonmetropolitan volume | 52.1% |
| Surgeons per 100,000: metro / nonmetro | 1.03 / 0.58 |
| Nonmetropolitan practice, OR for R/P pathway (95% CI) | 3.86 (2.62-5.68) |
| **R/P-only counties** (no FT surgeon) | **223** — 23.4M residents; 4.9M beneficiaries; 44.8% nonmetro |
| R/P-only counties vs FT-served counties | More rural (SMD +0.67), lower income (−0.47), more whole-county HPSA (+0.56) |
| R/P-only counties with ≥1 MDS-certified surgeon | 161/223 (72.2%) — 17.9M residents |
| Sole-provider counties; sole surgeon R/P-trained | 274; 162 (59.1%) |
| — of which sole surgeon is MDS-certified | 107 (66.0%) — 9.4M residents |
| R/P-only hospital service areas (HSAs) | 309/1,116 — 35.1M residents |
| Hospital referral regions with no FT surgeon | 28/298 — 9.8M residents |
| Certified surgeons: nonmetro share, R/P vs FT | 10.3% vs 7.0% (1.5×) |

Adjusted estimates under the training-based definition: Table VI. RUCC, Rural-Urban Continuum Code; HPSA, primary-care Health Professional Shortage Area; SMD, standardized mean difference; OR, odds ratio.

Seventy-one percent of R/P-trained surgeons describe themselves to Medicare as general dermatologists, and MMS accounts for a median 5.3% of their Medicare services, against 27.9% among FT surgeons (Supplementary Material S-M1), so rural Mohs surgery is largely a service line within full-scope rural dermatology practices. Certification does not reverse the geography. R/P-trained diplomates were 1.5 times as likely as FT diplomates to practice nonmetropolitan (10.3% vs 7.0%), and in RUCC 8-9 every certified Mohs surgeon was R/P-trained.

In 223 counties (44.8% of them nonmetropolitan), home to 23.4 million residents and 4.9 million Medicare beneficiaries, every Mohs surgeon was R/P-trained. These counties were more rural, poorer, and more often whole-county primary-care shortage areas than FT-served counties, and 28 entire Hospital Referral Regions (9.8 million residents) had no FT surgeon. Certification reached most of this geography, since 161 of the 223 counties (72.2%; 17.9 million residents) were served by at least one MDS-certified surgeon. In 107 of these counties (9.4 million residents), the only Mohs surgeon was a board-certified R/P-trained surgeon.

### Access under simulated exclusion (Table III; Figures 3-4)

With the R/P-trained workforce removed, the population-weighted mean drive time to the nearest Mohs practice rose from 17.6 to 24.3 minutes. The population beyond 60 minutes nearly doubled, from 17.2 to 32.8 million, and Medicare beneficiaries beyond 60 minutes rose from 4.3 to 7.9 million. Measured against the dermatology time-and-distance standards CMS applies to Medicare Advantage networks, the population outside the standards also nearly doubled, from 23.7 to 46.8 million, a difference of 23.1 million people, 5.2 million of them beneficiaries.

**Figure 3.**
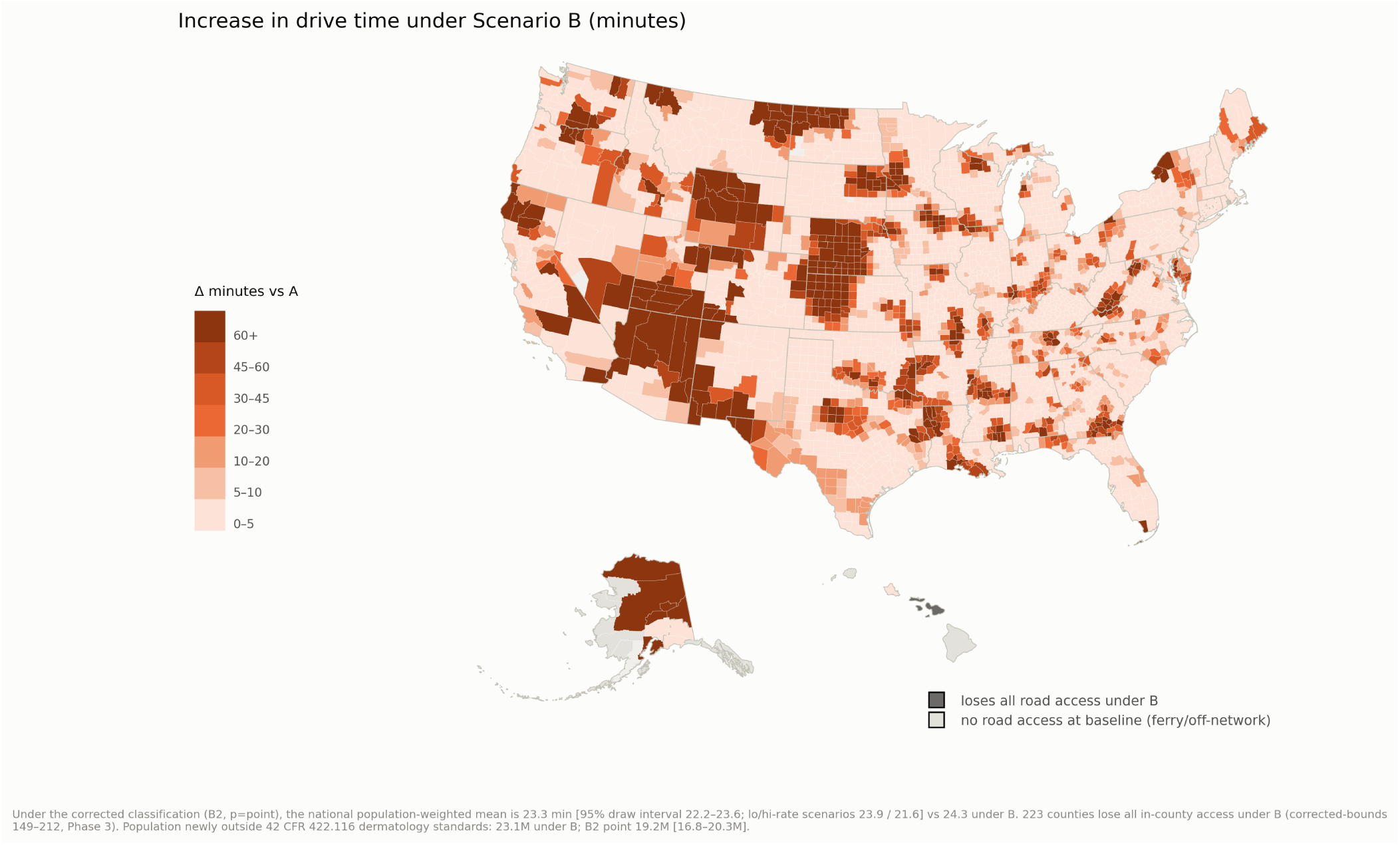
Change in drive time to the nearest Mohs surgeon under simulated exclusion. Increase in road-network drive time from populated-ZCTA population-weighted centroids, scenario B versus A. Status-quo and post-exclusion absolute maps: Supplementary Figures S1-S2.

**Figure 4.**
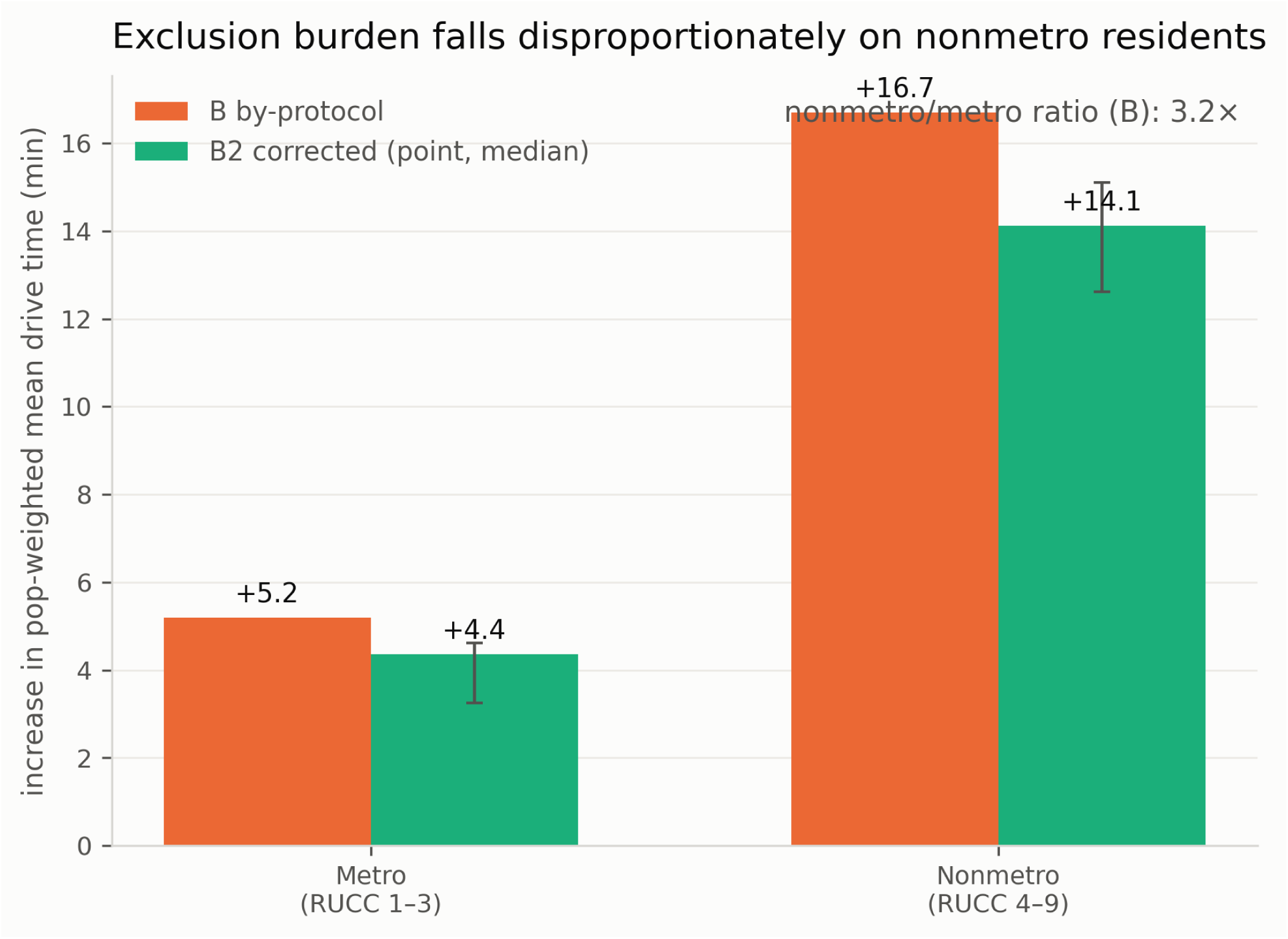
Rural-urban disproportionality of simulated access loss. Population-weighted mean drive-time increase and share of population newly beyond 60 minutes, metropolitan versus nonmetropolitan (scenario B; adjusted classification in Table III).

**Table III.** Simulated exclusion of R/P-trained surgeons: drive time and network adequacy (50 states + DC)

| Outcome | Status quo | Exclusion |
| --- | --- | --- |
| Population-weighted mean drive time, min | 17.6 | 24.3 (+6.8) |
| Population-weighted median, min | 9.9 | 12.2 |
| Population >60 min (millions) | 17.2 | 32.8 |
| Population >90 min (millions) | 6.1 | 14.9 |
| Population >120 min (millions) | 2.5 | 7.8 |
| Beneficiaries >60 min (millions) | 4.3 | 7.9 |
| Outside CMS MA dermatology time-and-distance standards (millions) | 23.7 | 46.8 |
| — newly noncompliant (millions) | — | 23.1 |
| — newly noncompliant beneficiaries (millions) | — | 5.2 |
| Counties losing all in-county access | — | 223 |
| Population losing all road-accessible access | — | 154,100 |
| Nonmetro vs metro mean increase | — | +16.7 vs +5.2 min (3.2×) |
| Nonmetro share of newly >60 min population | — | 47.7% (vs 14% of population) |
| Excluded annual Medicare volume (cases) | — | 353,736 |
| Unabsorbed volume if every surviving FT surgeon expands caseload 20% (0%/50%) | — | 211,848, 59.9% (353,736 / 94,982) |

Drive times: road-network (OSRM) from 26,419 populated-ZCTA population-weighted centroids; 466,000 people with no road access to any site at baseline reported separately. CMS MA standards: 42 CFR 422.116 dermatology maxima by county designation. Adjusted estimates: Table VI.

These losses were not evenly spread. All 223 R/P-only counties lost in-county access, in 186 of them the nearest remaining surgeon was out of state, and 154,100 Hawai’i residents lost road-accessible access altogether. Nonmetropolitan residents are 14% of the national population but were 47.7% of the people newly pushed beyond 60 minutes. The nonmetropolitan mean increase (+16.7 minutes) was 3.2 times the metropolitan one (+5.2). In the hardest-hit states, Alaska’s mean drive time rose from 34 to 129 minutes, Wyoming’s from 63 to 122, and New Mexico’s from 58 to 94.

### Distribution and concentration of the loss (Figure 6)

The loss is a tail phenomenon, and the mean increase of 6.8 minutes conceals it. For 61.8% of Americans the exclusion changes nothing, because their nearest surgeon is already fellowship-trained. The 95th percentile of drive time, however, moves from 61 to 86 minutes, the 99th from 110 to 177, and the population beyond two hours triples, from 2.5 to 7.8 million. The 20.6 million people facing increases above 30 minutes are 6.2% of the population, and they absorb 71% of the added travel time. Nor is the tail purely rural. About 6.0 million people in 322 ZCTAs whose entire local Mohs supply is R/P-trained would see the drive to the nearest practice jump from under 15 minutes to over 60, and those areas include metropolitan Bakersfield, Lancaster-Palmdale, and the west side of El Paso.

### Capacity and waiting

The excluded surgeons’ 353,736 annual Medicare cases would not disappear. They would join the schedules of the remaining workforce, and absorbing them would require the average remaining FT surgeon to increase annual volume by 45.5%. Two constraints stand in the way. The first is aggregate capacity, since even if every surviving FT surgeon increased caseload by 20%, 211,848 cases (59.9%) would go unabsorbed. The second is geography, which higher caseloads cannot overcome. Even at 50% more cases per surgeon, when national spare capacity would exceed the displaced cases, 94,982 cases (26.9%) would still have no surgeon within reach, because the spare capacity is not where the displaced patients are. Because 88.1% of the excluded volume is billed by surgeons who practice only in metropolitan counties, waits would lengthen in cities as well as the countryside, and the cases beyond any surgeon’s reach would surface as longer travel, substitution of non-Mohs treatment, or forgone care.

### Replacement capacity (Table IV; Figure 5)

The MSDO pipeline is capped by supply rather than by demand, since 88 accredited programs filled 99.2% of their positions from 2019 through 2024 and produce roughly 91-98 US-bound graduates per year. The constraint is where the graduates practice. Only 10.3% of the FT entrants of 2021-2024 added a nonmetropolitan practice site, so the two pathways contributed nearly equal numbers of new rural surgeons in those years, 37 FT and 34 R/P-trained. A fellowship-only restriction would therefore eliminate the source of about 48% of new rural Mohs capacity in addition to removing the majority of the rural surgeons already in practice. Where rural access is most fragile, the R/P pathway is the only observed source of new surgeons, since no FT surgeon began practice in any incumbent R/P-only county in 2021-2024, against 39 R/P-trained entrants who did. The replacement model’s base case takes the pipeline’s 2019-2025 pace (about 95 US-bound graduates per year), 10.3% of graduates adding a nonmetropolitan site, each rural arrival practicing at the mature rural-FT volume, and the exit rates measured in the 2021-2023 panel. The optimistic and pessimistic cases move graduates to 101 and 85 and rural siting to 12% and 6.7%. The optimistic siting rate is one step beyond the rising cohort trend, and the pessimistic rate is the nonmetropolitan share of the whole FT workforce (Supplementary Material S-M6). This headcount measure counts rural surgeons wherever in nonmetropolitan America they practice, so it is the measure more favorable to a restriction. Even so, the 2026 rural headcount is not regained for about 21 years after exclusion, 14 under the optimistic case and more than 30 under the pessimistic one, and rural capacity in 2041 remains roughly 39% below its no-restriction trajectory. Restoring the headcount within 10 years would require 18% of graduates to settle nonmetropolitan, 1.7 times the observed rate, and within 5 years 34%, a rate no graduating cohort has approached. In the counties themselves the loss is not recovered at all within the projection horizon, because no fellowship-trained surgeon entered any incumbent R/P-only county in 2021-2024 and the model therefore has no observed channel through which a fellowship graduate replaces an excluded surgeon there. Immediate exclusion ends in-county access in all 223 counties at once. A restriction with grandfathering avoids the immediate shock but not the outcome. Under grandfathering, the rural workforce in 2041 sits 24% below its no-restriction trajectory, and 114 of the 223 counties (range 96-130), home to 10.8 million residents, lack in-county access by 2041 as their surgeons retire unreplaced, against 77 (range 63-92) with no restriction (Figure 5, C). Retirement at a fixed age of 65 in place of measured exit rates gives the same restoration time (21 years) and smaller county counts, 73 (range 71-75) under grandfathering against 51 (range 44-58) with no restriction (Figure 5, dotted lines; Supplementary Material S-M6).

**Figure 5.**
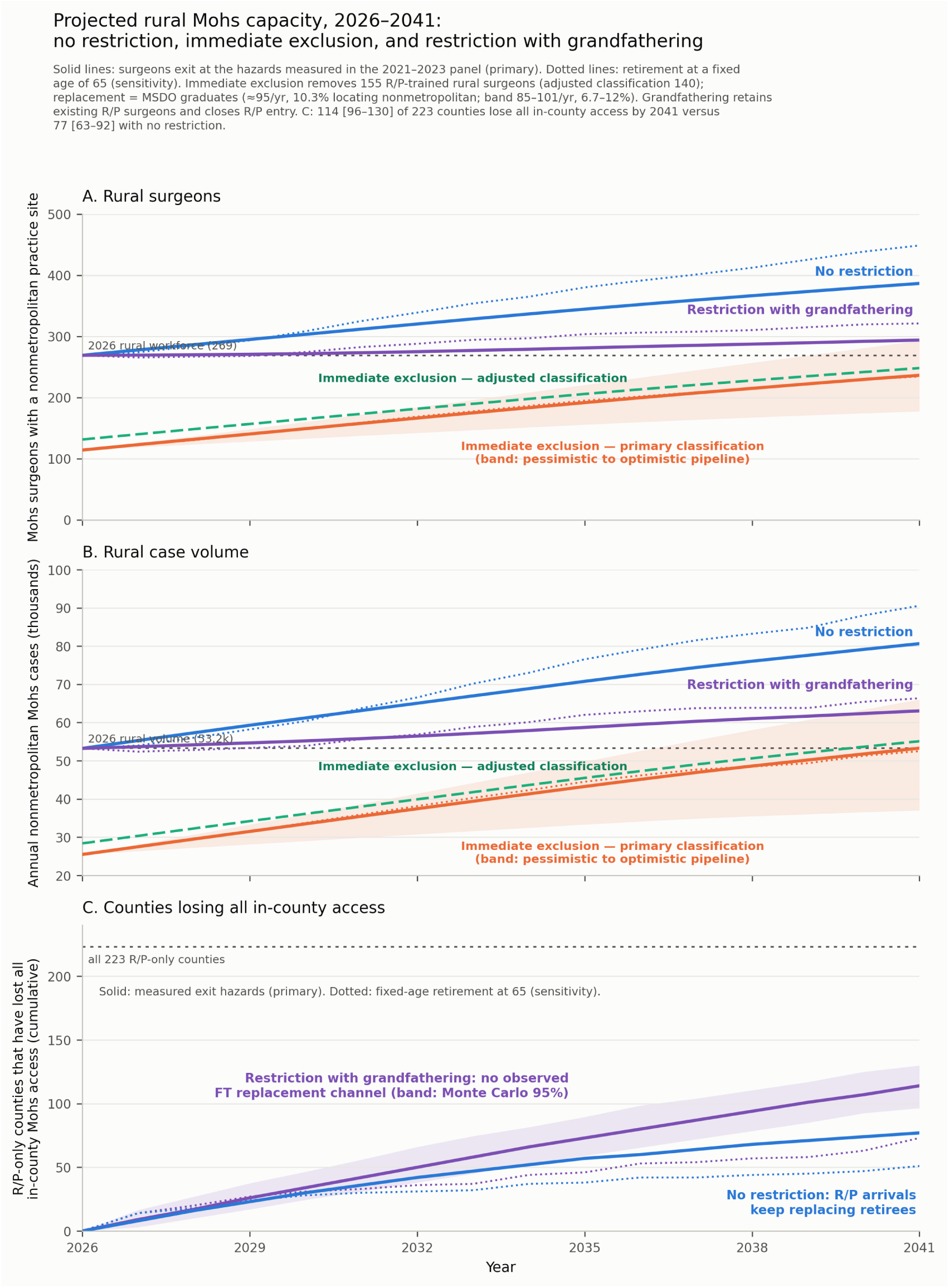
Projected rural Mohs capacity under no restriction, immediate exclusion, and a restriction with grandfathering, 2026-2041. One cohort model for all scenarios. A, Rural surgeon headcount, and B, annual nonmetropolitan case volume, under no restriction, immediate exclusion (primary and adjusted classification), and a restriction with grandfathering (existing R/P-trained surgeons retained; R/P entry closed). C, Cumulative R/P-only counties losing all in-county access, 114 [96-130] of 223 by 2041 under grandfathering versus 77 [63-92] with no restriction. Solid lines: surgeons exit at the hazards measured in the 2021-2023 panel (primary analysis); dotted lines: retirement at a fixed age of 65 (sensitivity analysis); dashed: adjusted classification; bands: pessimistic to optimistic pipeline assumptions (A, B) and the Monte Carlo 95% interval (C).

**Figure 6.**
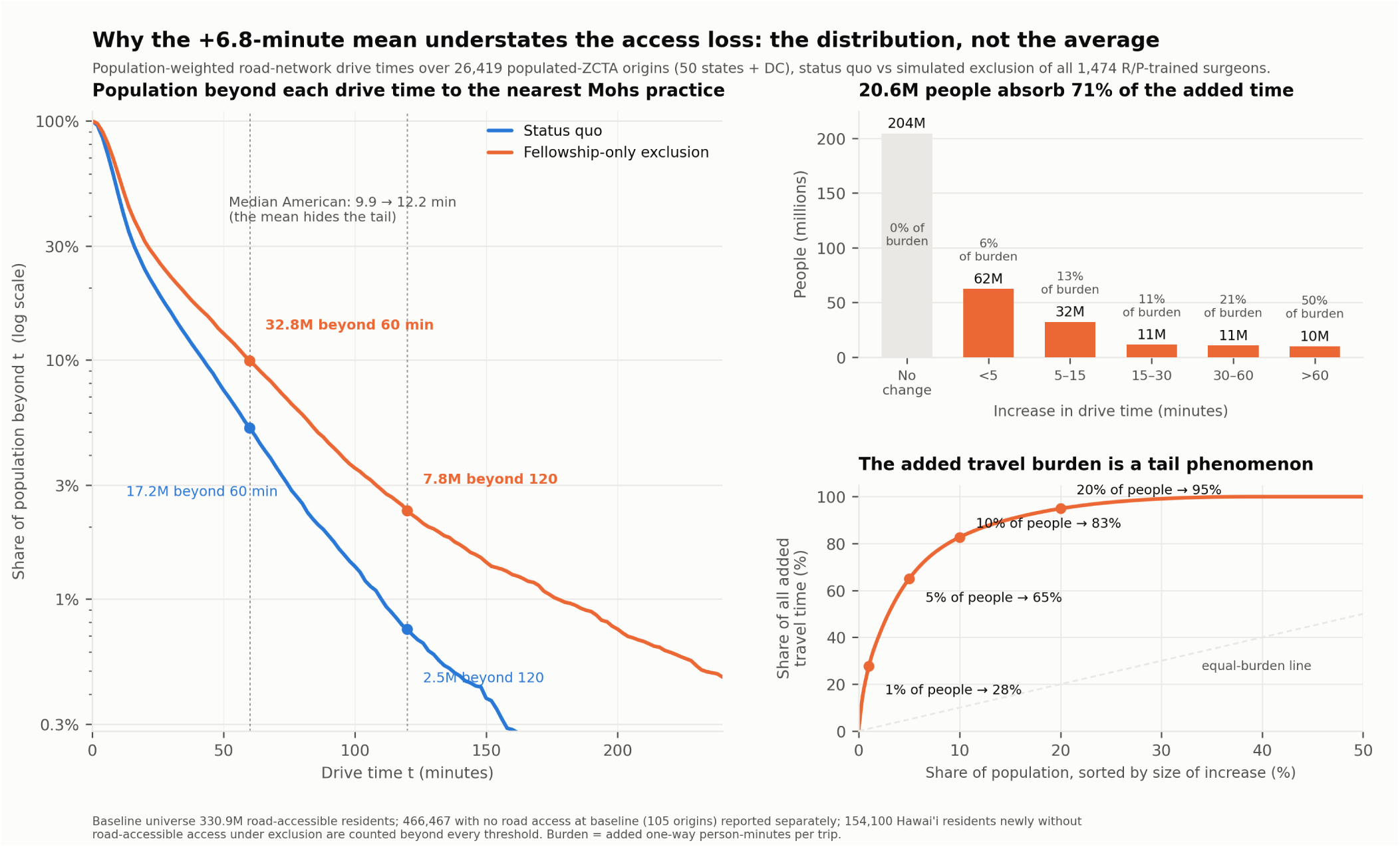
Distribution and concentration of the drive-time loss under simulated exclusion. Left, share of the population beyond each drive time, status quo versus exclusion (log scale). Right top, population by size of individual drive-time increase (61.8% see no change; the 20.6 million people with increases above 30 minutes absorb 71% of all added travel time). Right bottom, cumulative concentration of added travel time (the most-affected 5% absorb 65%). Universe 330.9 million road-accessible residents; 466,467 people (105 origins) with no road access at baseline are reported separately, and 154,100 Hawai’i residents newly without road access are counted beyond every threshold.

**Table IV.**
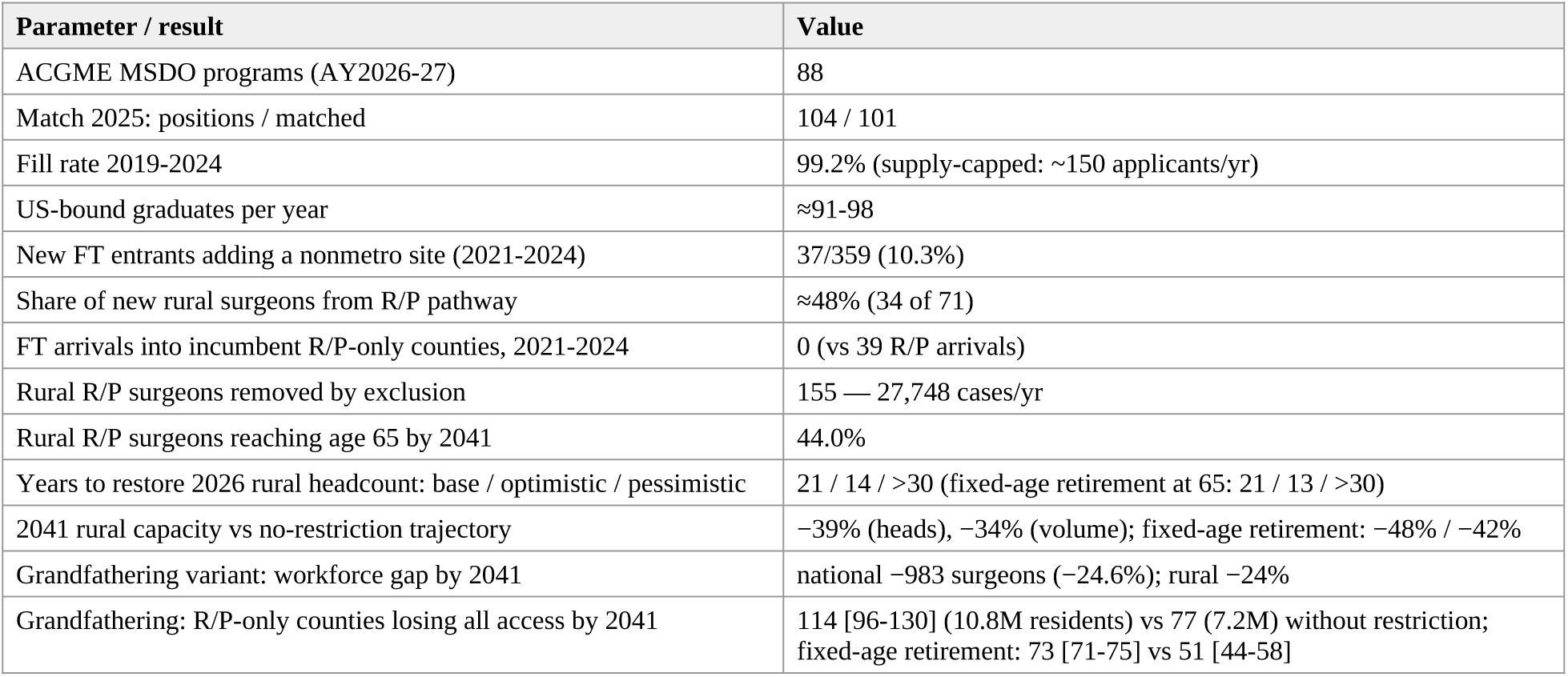
Fellowship pipeline and replacement of excluded rural capacity. Assumptions and sensitivity ranges in Supplementary Material; all pipeline figures from primary sources (ACGME, SF Match, ACMS match reports).

| Parameter / result | Value |
| --- | --- |
| ACGME MSDO programs (AY2026-27) | 88 |
| Match 2025: positions / matched | 104 / 101 |
| Fill rate 2019-2024 | 99.2% (supply-capped: ~150 applicants/yr) |
| US-bound graduates per year | ≈91-98 |
| New FT entrants adding a nonmetro site (2021-2024) | 37/359 (10.3%) |
| Share of new rural surgeons from R/P pathway | ≈48% (34 of 71) |
| FT arrivals into incumbent R/P-only counties, 2021-2024 | 0 (vs 39 R/P arrivals) |
| Rural R/P surgeons removed by exclusion | 155 — 27,748 cases/yr |
| Rural R/P surgeons reaching age 65 by 2041 | 44.0% |
| Years to restore 2026 rural headcount: base / optimistic / pessimistic | 21 / 14 / >30 (fixed-age retirement at 65: 21 / 13 / >30) |
| 2041 rural capacity vs no-restriction trajectory | –39% (heads), –34% (volume); fixed-age retirement: –48% / –42% |
| Grandfathering variant: workforce gap by 2041 | national –983 surgeons (–24.6%); rural –24% |
| Grandfathering: R/P-only counties losing all access by 2041 | 114 [96-130] (10.8M residents) vs 77 (7.2M) without restriction; fixed-age retirement: 73 [71-75] vs 51 [44-58] |

### Need for rural Mohs capacity (**Table VII**; **Figure 7**)

Rural access is short today. Of the 3,144 counties, 2,310 have no Mohs surgeon, among them 1,762 of the 1,958 nonmetropolitan counties (90%). Nationally, 23.7 million people live outside the CMS dermatology time-and-distance standard, 12.2 million of them in nonmetropolitan counties (3.1 million beneficiaries, in 975 counties). Bringing those beneficiaries inside the standard would take 63,800 more first-stage cases a year delivered within reach, which we call the case shortfall of 2026, the unmet need of today. The corresponding surgeon shortfall is 323 rural Mohs surgeons at the current rural caseload, more than the 269 who practice in nonmetropolitan counties today. Rural demand will not stay at its 2024 level, as the replacement model assumes, because rural residents aged 75-84 are projected to increase 18% by 2041 and those 85 and over 48%, and skin-cancer treatment per beneficiary grew 3.2% a year over 2006-2012. Need therefore reaches 202,000 rural first-stage cases a year in 2041, 1.7 times the 2026 need, or 1,021 rural Mohs surgeons, and every scenario falls short of need from 2026 on (Figure 7). With no restriction, rural capacity reaches about 386 surgeons and 80,600 cases a year in 2041, leaving a surgeon deficit of 634 and a case deficit of 121,000 a year. Under a restriction with grandfathering the deficits are 727 surgeons and 139,000 cases, and under immediate exclusion 784 and 149,000. Meeting need by 2041 would take about 77 new rural Mohs surgeons a year from both pathways, against about 18 today, and even closing today’s gap alone, with no growth in Mohs use per beneficiary, would take about 41 a year. The fellowship pipeline alone would have to place 55% of a 95-graduate class in rural practice, or 58% with the R/P workforce removed as well. Status quo access alone takes 464 surgeons in 2041 and about 24 entrants a year. Measured against that target, the pipeline as it stands keeps pace only if Mohs use per beneficiary grows less than 2.3% a year, a restriction with grandfathering only if it grows less than 0.6%, and immediate exclusion at no rate at all. The sensitivities of Table VII put the entry required between 34 and 118 a year. The lower figure counts in the gap only the 2.1 million nonmetropolitan residents more than two hours from a Mohs surgeon (a case shortfall of 12,500 today), and the upper applies the unadjusted fee-for-service trend. The population outside the standard rises from 23.7 million today to 28.2 million by 2041 with no restriction, 30.9 million under a restriction with grandfathering, and 46.8-47.6 million under immediate exclusion (Figure 7, C).

**Figure 7.**
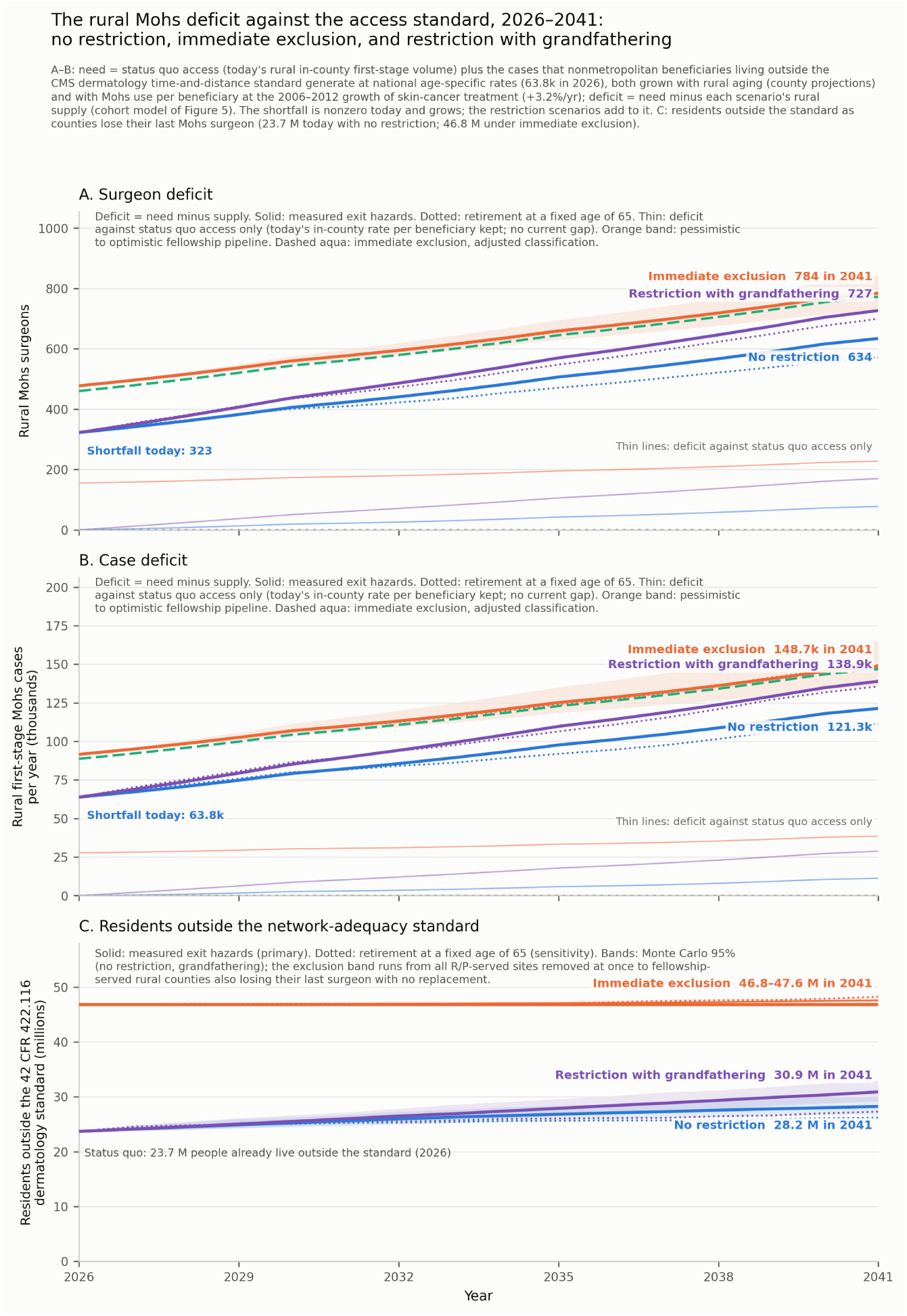
The rural Mohs deficit against the access standard under no restriction, immediate exclusion, and a restriction with grandfathering, 2026-2041. A, Surgeon deficit, and B, case deficit: rural Mohs surgeons and annual rural first-stage Mohs cases short of need, the capacity that keeps status quo access and brings nonmetropolitan beneficiaries inside the CMS dermatology time-and-distance standard as rural beneficiaries age and Mohs use grows at the 2006-2012 rate of skin-cancer treatment (need minus supply; Table VII). The shortfall today is 323 surgeons and 63,800 cases a year; by 2041 the deficit is 634 surgeons and 121,000 cases with no restriction, 727 and 139,000 under a restriction with grandfathering, and 784 and 149,000 under immediate exclusion. Each scenario is drawn twice in its own color, solid for the measured exit hazards (primary analysis) and dotted for retirement at a fixed age of 65 (sensitivity analysis); the thin lines near the bottom of each panel are the same deficits measured against status quo access alone; the dashed green line is immediate exclusion under the adjusted classification; the orange band spans pessimistic to optimistic pipeline assumptions. C, Residents living outside the standard (42 CFR 422.116) as counties lose their last Mohs surgeon: 23.7 million today; 28.2 [26.8-30.2] million by 2041 with no restriction, 30.9 [29.2-33.0] million under a restriction with grandfathering, and 46.8-47.6 million under immediate exclusion. Solid and dotted lines as in A and B; bands are the Monte Carlo 95% interval (no restriction, grandfathering) and, for exclusion, the range from all R/P-served sites removed at once to fellowship-served rural counties also losing their last surgeon without replacement.

### Results under a training-based (adjusted) definition of fellowship training (Table VI)

Directory listing is a membership behavior, so a fellowship-trained surgeon who is not a member is misclassified R/P-trained. The seeded audit found publicly documented fellowship-training claims in 3 of 30 randomly sampled R/P-classified surgeons (10.0%; Wilson 95% CI 3.5%-25.6%), and every principal result was therefore re-derived with non-audited R/P-classified surgeons reclassified FT by fixed-seed Monte Carlo at the audited rate and at its Wilson limits (Table VI). The adjustments are modest and directionally consistent, and no conclusion changes at any point in the range, including the extreme upper bound. Even at 25.6% misclassification, 149 counties retain no fellowship-trained surgeon, 13.4 million people fall newly outside CMS standards, and grandfathering still leaves 81 of the remaining R/P-only counties without in-county access by 2041.

## DISCUSSION

A fellowship-only restriction on Mohs surgery would produce a large loss of patient access, concentrated in rural counties, and the fellowship pipeline could not repair the loss. Five findings support that conclusion. First, R/P-trained surgeons are a structural pillar of American Mohs surgery, not a periphery: they are 45% of the active workforce, most of them are subspecialty board-certified, and they deliver the majority of nonmetropolitan Mohs care. Second, removing them would produce a large and geographically concentrated loss of access, the unintended consequence that any body contemplating a training-based standard would have to weigh. Twenty-three million people would find themselves newly outside the dermatology travel standards CMS applies to Medicare Advantage networks, nearly doubling the population already outside those standards (23.7 to 46.8 million). The losses would fall three times as heavily on nonmetropolitan America, and in 223 counties access would end outright.

Third, the loss is not replaced. In aggregate, the pipeline would take about two decades to rebuild the rural headcount, and would have to send a third of every graduating class to nonmetropolitan practice over a period of five years. In the counties that lose their surgeons, access is not rebuilt at all, because the only observed replacement channel into those counties is the R/P pathway. Even grandfathering leaves 114 of those counties stranded by 2041 due to attrition. Fourth, certification and fellowship are distinct populations, and a fellowship-only rule would exclude 955 board-certified surgeons, including the sole, certified Mohs surgeon of 107 counties. Finally, rural access is already short of the CMS standard by 323 surgeons, an unmet need today, and meeting that need through 2041 would take about four times the rural entry rate both pathways now supply.

Beyond travel, the other consequence is waiting, and waiting is how an access loss that looks rural on a map reaches urban patients, since 88% of the displaced cases are metropolitan and land on metropolitan schedules. Dermatology already posts among the longest appointment waits of any specialty tracked in the national survey literature, 36.5 days on average in 2025 in 15 large metropolitan markets.^42^ A restriction that displaces roughly 354,000 Medicare cases per year onto a workforce that cannot absorb 60% of them would therefore lengthen waits everywhere. In skin cancer, waiting is not neutral, since surgical delays of under one year are associated with tumor growth in moderately and poorly differentiated squamous cell carcinoma,^43^ and longer treatment delay is associated with larger surgical defects.^44^ We did not model appointment availability, because claims data cannot see scheduling, but the capacity arithmetic establishes the direction, and metropolitan and rural patients alike would wait longer.

The findings answer questions that prior national analyses did not pose (Table V). Those analyses described the workforce’s geography,^19^ its society affiliations,^9^ its certification,^10^ and its decade trends^45^ without asking how access depends on training pathway. Our results come from the same public sources, with marginals matching those studies, and the answers are unambiguous. R/P-trained surgeons are the backbone of Mohs access in nonmetropolitan and rural America and a critical component of the national Mohs workforce. A fellowship-only restriction would fall disproportionately on rural America and, amid a skin-cancer epidemic, would cause access losses that are severe everywhere and total in 223 counties. The policy task is to strengthen that workforce, not to remove it.

**Table V.** Questions unaddressed in prior national Mohs workforce analyses, and this study’s findings.

| Question | Prior published analyses | This study |
| --- | --- | --- |
| What share of rural Mohs care does each training pathway deliver? | Workforce reported as 94.6% metropolitan without pathway stratification<sup>19</sup>; nonmetro shares tabulated by society membership (ACMS 3.3% vs ASMS 10.5% vs unaffiliated 8.1%) without statistical testing or discussion<sup>9</sup>; pathway and rurality fields co-present but not cross-tabulated<sup>10</sup>; after this analysis was completed, an independent 2023 cross-section (ZIP-code commuting-area rurality) reported 70.4% of rural Mohs billers not ACMS-listed and rural density falling from 0.44 to 0.13 per 100,000 if only listed surgeons are counted, but did not examine which counties would be left with no Mohs surgeon, patient travel time, surgeon certification, or the replacement of surgeons lost to retirement and other attrition<sup>25</sup> | R/P-trained surgeons are 57.6% of nonmetro surgeons and deliver 52.1% of nonmetro volume; FT share falls 58.4% → 0 across RUCC 1 → 9; nonmetro OR for R/P 3.86 (adjusted 3.29) |
| Where do board-certified surgeons without fellowship training practice? | Pathway composition of diplomates reported (27-33% non-fellowship)<sup>58,59</sup>; geography not shown<sup>10</sup> | 1.5× as likely as FT diplomates to practice nonmetro (10.3% vs 7.0%); in RUCC 8-9 all certified surgeons are R/P-trained; 72.2% of |
|  |  | R/P-only counties have a certified surgeon |
| What would excluding R/P-trained surgeons do to patient access? | Not previously simulated; county-level access described without pathway overlay<sup>20,22</sup>; trends analyzed at state level<sup>45</sup> | +6.8 min mean drive time; 15.6M more people beyond 60 min; 23.1M newly outside CMS MA standards; 223 counties lose all access; 3.2× rural disproportionality |
| Could the fellowship pipeline replace excluded rural capacity? | Graduate supply and metropolitan location documented (96% metro)<sup>24</sup>; adequacy against a restriction not modeled | ≈21 years to restore rural headcount (base case); zero observed FT entry into incumbent R/P-only counties 2021-2024; grandfathering still strands 114 counties by 2041 |
| Do clinical outcomes differ by training pathway? | No comparative outcome study exists in either direction (systematic-style search); VHA analysis found no stages-per-case difference by ACMS membership (P=.59)<sup>8</sup> | Not measurable in claims; parity on the specialty's own quality measure (median 1.5 stages/case in both pathways)<sup>41</sup> — a proxy for surgical judgment and appropriate practice, not a measured clinical outcome |

One objection raised in that literature deserves a direct answer, namely that many diplomates certified through the practice pathway performed few Medicare Mohs cases in the year examined and so should not be counted as real capacity.^10^ The objection measures a billing window rather than a practice, because fee-for-service claims see only part of a surgeon’s work, and Medicare Advantage, now roughly half of Medicare enrollment, and all non-Medicare care lie outside those claims. From the patient’s side the objection cuts the other way, since a surgeon who bills little may be a community’s only Mohs surgeon. Among diplomates billing fewer than 50 Medicare cases, 17 were the only Mohs surgeon in their county, and those 17 counties hold 2.9 million residents.

A fellowship-only restriction rests on the premise that fellowship-trained surgeons produce better outcomes than residency- and practice-trained surgeons. No comparative outcome study of the two pathways exists, in either direction. The outcome literature on Mohs surgery, on complication rates, cure rates against excision, and frozen-section accuracy, describes the procedure without regard to the surgeon’s training route,^46–49^ and the two clinical series of surgeons trained outside fellowship report rates within the published range but have no fellowship-trained comparison group.^5,7^ The pathways have been compared only on stages per case, the specialty’s standard quality measure and a proxy for surgical judgment, and on that proxy they are indistinguishable. The medians are 1.5 and 1.5 here, and the VHA, the one system that has implemented a fellowship-based restriction, found 1.52 vs 1.57 between fellowship-trained (ACMS-member) surgeons and others in its first nationwide assessment (149 surgeons, 2017-2024; P=.59).^8^ The only proxy finding in the fellowship direction is a single-year outlier association by ACMS membership,^50^ while the three-year persistence analysis of the same measure found none^41^ and outliers occur on both sides of the membership line.^51^ The burden of proof lies with the proponents of a restriction to show that its benefits outweigh its harms. The harms are measured here; no study has shown a clinically meaningful benefit.

Current federal policy already embodies the appropriate pathway-neutral standard. Under CLIA’s personnel rules as CMS now applies them, board-certified dermatologists, whether certified by the ABD or the American Osteopathic Board of Dermatology (AOBD), are qualified to direct laboratories performing dermatopathology testing, including Mohs frozen sections, and fellowship training is not required.^12–15^ The specialty asked CMS for that standard, through the certifying boards, the American Academy of Dermatology (AAD), the ACMS, the American Society for Dermatologic Surgery Association (ASDSA), and the ASMS,^52–56^ and Medicare coverage policy recognizes fellowship, residency, course, and preceptorship training in Mohs surgery alike.^57^ Many R/P-trained surgeons (55.6%) also hold the MDS subspecialty certification, but since 2026 that examination has been open only to fellowship graduates, the practice pathway having closed after 2025 under the customary sunset for a new subspecialty certificate. Nothing in that sunset speaks to competence, since 955 R/P-trained surgeons passed the same examination while the pathway was open. Laboratory directorship should therefore stay keyed to primary dermatology board certification, the credential under which dermatologists have directed these laboratories for decades.

These estimates attach to the exclusion itself rather than to any single instrument. Practice-level instruments such as privileging standards and laboratory-director qualifications remove the practice for all payers, so the population-denominated results apply, whereas payer-level instruments such as network credentialing or payment rules tied to CPT 17311-17315 exclude only the covered population, so the beneficiary-denominated results are the prediction for Medicare. The VHA directive is an implemented member of this class but not a template for the others. It restricts privileging inside a salaried system in which the Mohs laboratory sits under the facility pathology service, so the excluded surgeon loses a privilege rather than the directorship on which a community practice depends, and it operates behind a purchased-care valve that Medicare fee-for-service lacks, Community Care referral at 60-minute and 28-day standards (38 CFR 17.4040). It has produced no demonstrated benefit^8^ while its cost to veterans’ access has gone unreported. In the community the directorship is the practice, and there is no Community Care program for the county that loses it.

### Policy implications

Even with no restriction at all, 77 of the 223 counties that depend solely on R/P-trained surgeons lose their last in-county Mohs surgeon by 2041 (Figure 5C), because arrivals (39 R/P-trained surgeons and no fellowship-trained surgeons in 2021-2024) barely keep pace with retirements. The status quo is a slow leak in rural access; a restriction would turn it into a rupture. Both call for the same remedy, more surgeons trained to practice Mohs surgery where patients live, through every route that produces them. The fellowship pipeline should grow, but at the observed rate of rural siting a larger pipeline adds only a few rural surgeons a year, and its observed flow into fellowship-absent counties has been zero. Meeting need by 2041 would take about 77 new rural Mohs surgeons a year, against about 18 arriving now from both pathways. The fellowship pipeline alone could supply that number only by placing more than half of every class in rural practice or by growing more than fivefold, and closing the R/P route would remove the only lever now operating at scale in the counties at issue. We recognize that many of the residents counted outside the standard are treated today, by traveling farther than the standard allows. Need as defined here is the capacity that would have to sit within reach for them to have the access the standard defines, and even when the gap counts only residents beyond a two-hour drive, a shortfall remains today and every scenario still misses need in 2041 (Table VII). Securing rural access therefore requires more routes into Mohs surgery, not fewer. Dermatology residencies should strengthen Mohs surgical training, as surgically robust programs already do, including in rural-track programs. The structured practice-based routes of courses, preceptorships, and proctored experience, which Medicare coverage policy already recognizes,^57^ should be sustained. And the certifying boards should reopen a practice-based route to the MDS examination, which certified 955 R/P-trained surgeons before it closed. For regulators and payers, the implication is to keep laboratory-director and network eligibility keyed to primary board certification and to weigh access explicitly before adding any training-based barrier. Fellowship is *one* valuable pathway, but not the *only* door.

### Limitations

The census sees only Medicare fee-for-service billers, so the 124 surgeons hidden by PUF suppression and all practice at the VA, in Kaiser-type integrated systems, and outside Medicare are invisible, and each of these gaps understates R/P-trained capacity and therefore the access losses reported here. Pathway classification uses the directory definition operative in prior national analyses, and its error runs in the opposite direction, because a fellowship-trained surgeon who is not a directory member is counted R/P-trained, a miscount that overstates the by-protocol exclusion impact. The adjusted results (Table VI) are lower, with no conclusion changed. The audit’s public-bio method can miss fellowship training for surgeons with sparse web presence, so the audited rate is itself a lower bound, and the net direction of these two opposing errors cannot be determined. Drive times assume free-flow speeds and exclude 466,000 people with no road access at baseline, who are reported separately. Appointment waiting times were not modeled, so the waiting-time implications rest on capacity arithmetic, and because the absorption scenarios use a prespecified caseload-expansion parameter that is assumed rather than estimated, the unabsorbed volume should be read as the 0-50% band (353,736 to 94,982 cases per year). The drive-time, network-adequacy, and county-access findings do not depend on that parameter. Exit hazards were measured over a three-year panel and held constant, and the fixed-age alternative appears in Supplementary Material S-M6. The need estimate rests on four inputs, and we have tried to be plain about each. It takes the CMS dermatology time-and-distance standard as the definition of adequate access, although Mohs surgery, which many patients travel farther for, has no standard of its own, and the two-hour sensitivity bounds that objection. It rests on county population projections. It carries a growth rate for Mohs use per beneficiary taken from the growth of all skin-cancer procedures in 2006-2012, below the rate at which Mohs cases per fee-for-service beneficiary rose over 2013-2024 even after adjustment for the shift of beneficiaries into Medicare Advantage. And it runs through a cohort model that holds cases per surgeon fixed although caseloads rose over 2020-2024, so that part of the need could be met by throughput rather than headcount (Table VII). The estimate’s age-specific rates measure the care delivered rather than clinical need. Finally, no clinical outcomes were measured. Stages per case is a proxy for surgical judgment and appropriate practice, and nothing in this study speaks to the clinical outcomes of care in either pathway.

**Table VI.** Principal results under the directory-based (primary) and training-based (adjusted) definitions.

| Result | Primary analysis (directory-based) | Adjusted, point [95% MC interval] | Adjusted bounds* |
| --- | --- | --- | --- |
| FT share of 2024 workforce | 54.7% | 59.3% | 56.3-66.3% |
| R/P share of nonmetropolitan volume | 52.1% | 47.3% [42.4-50.3] | — |
| FT share, nonmetropolitan surgeons | 42.4% | 48.0% [45.4-50.6] | 44.2-56.9% |
| Nonmetropolitan practice, OR for R/P | 3.86 (2.62-5.68) | 3.29 (2.67-3.97) | — |
| Counties with no FT surgeon | 223 (23.4M residents) | 192 [178-201] (19.5M) | 149-212 |
| — with ≥1 MDS-certified surgeon | 72.2% | 71.1% | 69.8-71.8% |
| R/P-only Hospital Service Areas | 309 (35.1M) | 262 [247-276] | 199-292 |
| Referral regions with no FT surgeon | 28 (9.8M) | 23 [19-27] | 16-26 |
| Mean drive time under exclusion (min) | 24.3 | 23.3 [22.2-23.6] | 21.6-23.9 |
| Population >60 min (millions) | 32.8 | 30.3 [28.1-31.3] | 26.3-31.9 |
| Newly outside CMS MA standards (millions) | 23.1 | 19.2 [16.8-20.3] | 13.4-21.6 |
| — beneficiaries (millions) | 5.2 | 4.3 [3.8-4.6] | 3.0-4.9 |
| Unabsorbed volume, +20% headroom (cases/yr) | 211,848 | 175,100 [166,286-185,698] | — |
| Rural surgeons removed | 155 | 140 [133-147] | — |
| Years to restore rural headcount (base case) | 21 | 19 | — |
| Grandfathering: counties losing access by 2041 | 114 [96-130] | 101 [86-117] | 81-109 |

\*Envelope of medians at the Wilson 95% limits (3.5%, 25.6%). MC, Monte Carlo (500 draws, fixed seed; 100 draws for routing-based metrics). Full machinery in Supplementary Material S-M2.

**Table VII.**
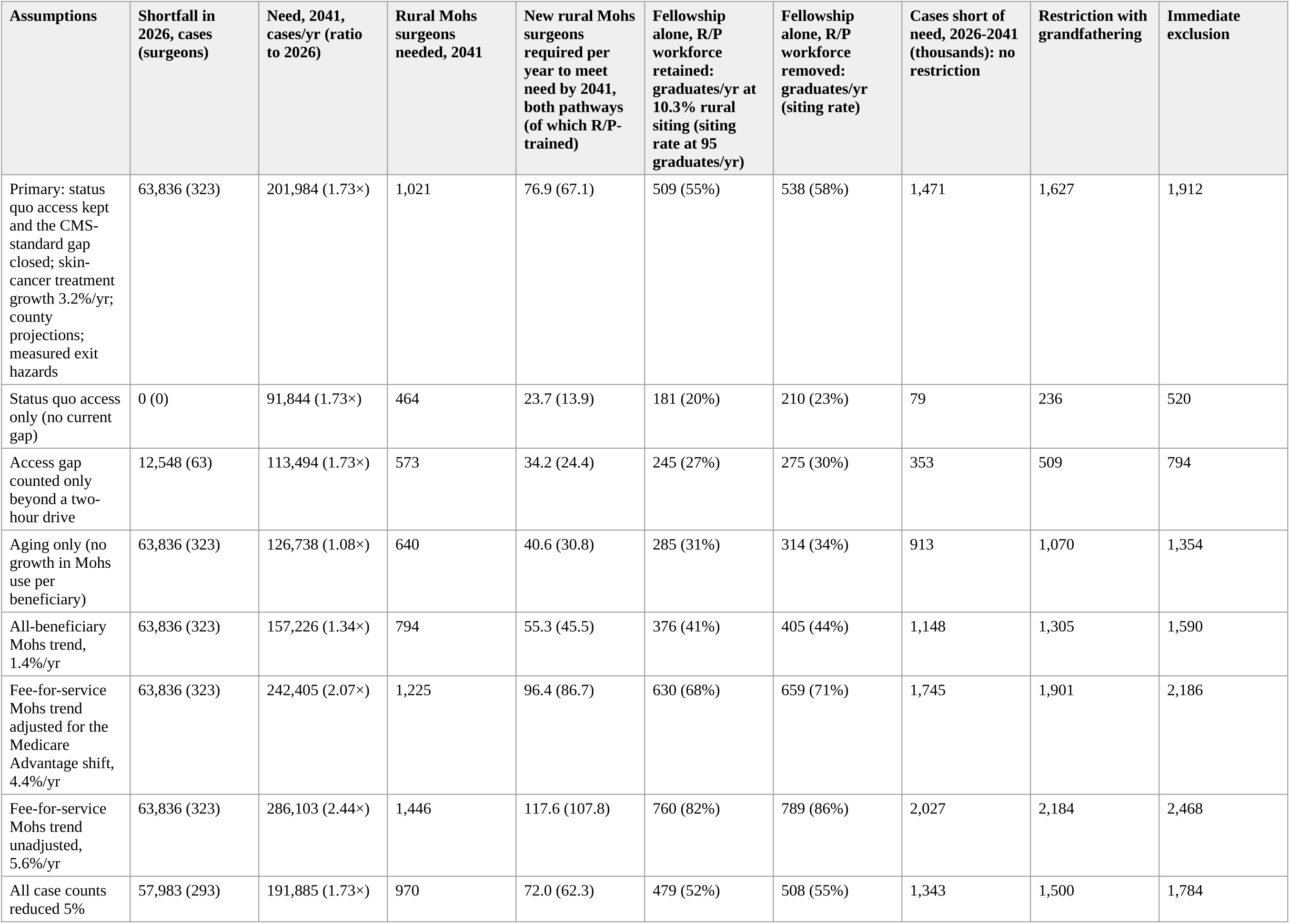
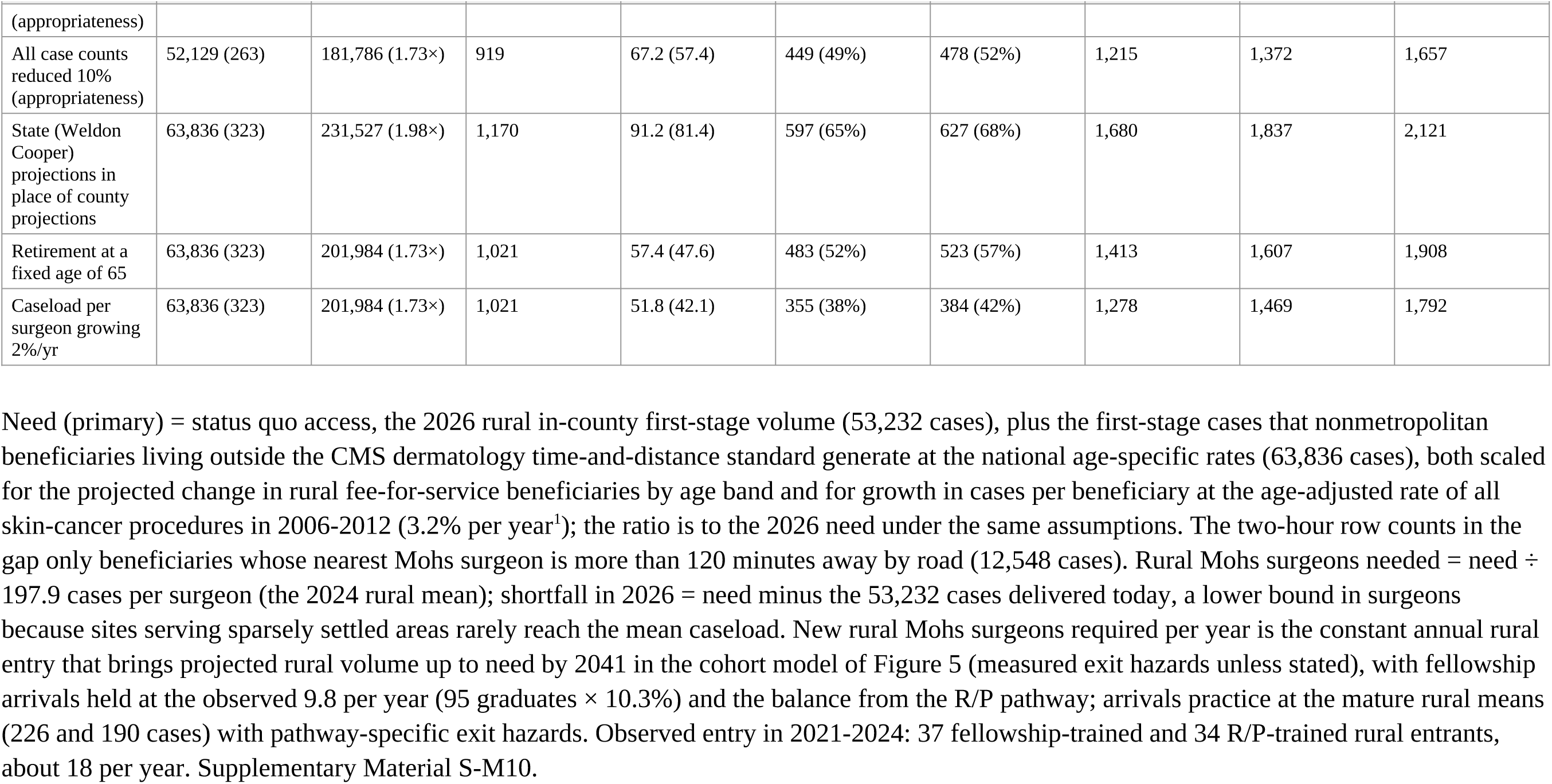
Rural Mohs capacity needed through 2041 to keep status quo access and bring rural residents inside the CMS access standard.

| Assumptions | Shortfall in 2026, cases (surgeons) | Need, 2041, cases/yr (ratio to 2026) | Rural Mohs surgeons needed, 2041 | New rural Mohs surgeons required per year to meet need by 2041, both pathways (of which R/P-trained) | Fellowship alone, R/P workforce retained: graduates/yr at 10.3% rural siting (siting rate at 95 graduates/yr) | Fellowship alone, R/P workforce removed: graduates/yr (siting rate) | Cases short of need, 2026-2041 (thousands): no restriction | Restriction with grandfathering | Immediate exclusion |
| --- | --- | --- | --- | --- | --- | --- | --- | --- | --- |
| Primary: status quo access kept and the CMS-standard gap closed; skin-cancer treatment growth 3.2%/yr; county projections; measured exit hazards | 63,836 (323) | 201,984 (1.73×) | 1,021 | 76.9 (67.1) | 509 (55%) | 538 (58%) | 1,471 | 1,627 | 1,912 |
| Status quo access only (no current gap) | 0 (0) | 91,844 (1.73×) | 464 | 23.7 (13.9) | 181 (20%) | 210 (23%) | 79 | 236 | 520 |
| Access gap counted only beyond a two-hour drive | 12,548 (63) | 113,494 (1.73×) | 573 | 34.2 (24.4) | 245 (27%) | 275 (30%) | 353 | 509 | 794 |
| Aging only (no growth in Mohs use per beneficiary) | 63,836 (323) | 126,738 (1.08×) | 640 | 40.6 (30.8) | 285 (31%) | 314 (34%) | 913 | 1,070 | 1,354 |
| All-beneficiary Mohs trend, 1.4%/yr | 63,836 (323) | 157,226 (1.34×) | 794 | 55.3 (45.5) | 376 (41%) | 405 (44%) | 1,148 | 1,305 | 1,590 |
| Fee-for-service Mohs trend adjusted for the Medicare Advantage shift, 4.4%/yr | 63,836 (323) | 242,405 (2.07×) | 1,225 | 96.4 (86.7) | 630 (68%) | 659 (71%) | 1,745 | 1,901 | 2,186 |
| Fee-for-service Mohs trend unadjusted, 5.6%/yr | 63,836 (323) | 286,103 (2.44×) | 1,446 | 117.6 (107.8) | 760 (82%) | 789 (86%) | 2,027 | 2,184 | 2,468 |
| All case counts reduced 5% | 57,983 (293) | 191,885 (1.73×) | 970 | 72.0 (62.3) | 479 (52%) | 508 (55%) | 1,343 | 1,500 | 1,784 |
| (appropriateness) |  |  |  |  |  |  |  |  |  |
| All case counts reduced 10% (appropriateness) | 52,129 (263) | 181,786 (1.73×) | 919 | 67.2 (57.4) | 449 (49%) | 478 (52%) | 1,215 | 1,372 | 1,657 |
| State (Weldon Cooper) projections in place of county projections | 63,836 (323) | 231,527 (1.98×) | 1,170 | 91.2 (81.4) | 597 (65%) | 627 (68%) | 1,680 | 1,837 | 2,121 |
| Retirement at a fixed age of 65 | 63,836 (323) | 201,984 (1.73×) | 1,021 | 57.4 (47.6) | 483 (52%) | 523 (57%) | 1,413 | 1,607 | 1,908 |
| Caseload per surgeon growing 2%/yr | 63,836 (323) | 201,984 (1.73×) | 1,021 | 51.8 (42.1) | 355 (38%) | 384 (42%) | 1,278 | 1,469 | 1,792 |

## CONCLUSION

Residency/practice-trained surgeons are a pillar of American Mohs surgery: 45% of the workforce, mostly board-certified, and the majority of its rural supply. Restricting MMS to fellowship-trained surgeons, through any instrument, would remove that pillar. It would push tens of millions of Americans, disproportionately rural, poorer, and shortage-designated, beyond CMS’s access standards, and the fellowship pipeline would not repair the loss, since it would take two decades to rebuild the rural headcount in aggregate and in four years has sent no graduate into the counties at issue. Residency- and practice-based training supplies the majority of the rural Mohs workforce, roughly half of its new capacity, and all observed entry into fellowship-absent counties. Even the status quo loses 77 of those counties by 2041, and keeping status quo access while closing today’s gap would take about four times the rural entry both pathways now supply. Securing rural access therefore means expanding every route into Mohs surgery, through more fellowship positions, stronger Mohs training within dermatology residency, practice-based training that certifying boards and payers continue to recognize, and a reopened practice pathway to MDS certification, while keeping regulatory eligibility keyed to primary dermatology board certification, which both pathways share. Patients need Mohs surgeons where they live. A fellowship-only restriction would remove surgeons from the communities that have the fewest and lengthen waits everywhere else. In a skin-cancer epidemic, all Americans deserve high-quality Mohs surgery within their reach, and securing it means strengthening both pathways, fellowship and residency/practice alike, and avoiding policies that would narrow access to care.

## ABBREVIATIONS USED

AAD: American Academy of Dermatology
ABD: American Board of Dermatology
ACGME: Accreditation Council for Graduate Medical Education
ACMS: American College of Mohs Surgery
AI: artificial intelligence
AOBD: American Osteopathic Board of Dermatology
ASDSA: American Society for Dermatologic Surgery Association
ASMS: American Society for Mohs Surgery
AY: academic year
CFR: Code of Federal Regulations
CI: confidence interval
CLIA: Clinical Laboratory Improvement Amendments
CMS: Centers for Medicare & Medicaid Services
CPT: Current Procedural Terminology
DC: District of Columbia
FT: fellowship-trained
HPSA: Health Professional Shortage Area
HRR: Hospital Referral Region
HSA: Hospital Service Area
IQR: interquartile range
MA: Medicare Advantage
MC: Monte Carlo
MDS: Micrographic Dermatologic Surgery
MMS: Mohs micrographic surgery
MSDO: Micrographic Surgery and Dermatologic Oncology
NPI: National Provider Identifier
OR: odds ratio
OSRM: Open Source Routing Machine
PUF: public use file
R/P-trained: residency/practice-trained
RUCC: Rural-Urban Continuum Code
SMD: standardized mean difference
STROBE: Strengthening the Reporting of Observational Studies in Epidemiology
USDA: US Department of Agriculture
VA: Department of Veterans Affairs
VHA: Veterans Health Administration
ZCTA: ZIP Code Tabulation Area

## DECLARATION OF GENERATIVE AI AND AI-ASSISTED TECHNOLOGIES IN THE MANUSCRIPT PREPARATION PROCESS

During the preparation of this work the authors used Claude (Anthropic) to assist with data analysis and with drafting and editing of the manuscript from the study’s tabulated outputs. After using this tool, the authors reviewed, edited, and verified the content, and take full responsibility for the content of the publication.

## Reprint requests

Kenneth K. Yu, MD, PhD, FAAD

## IRB approval status

Not applicable. The study used exclusively publicly available administrative and directory data on physicians; no patient-level data were used.

## Patient consent

Not applicable; the study contains no patient images or identifiable patient information.

## Clinical trial registration

Not applicable.

## Prior publication

None of the data have been published previously.

## Data availability

This study used only publicly available data; no new data were collected. The primary sources are the CMS Medicare Physician & Other Practitioners Public Use Files (by Provider and Service, and by Provider), NPPES, the American College of Mohs Surgery and American Society for Mohs Surgery public directories, American Board of Dermatology public diplomate lists, the CMS Doctors and Clinicians file, the CMS Medicare Monthly Enrollment and Physician & Other Practitioners by Geography and Service datasets, USDA ERS Rural-Urban Continuum Codes, U.S. Census geography and population estimates, published county population projections by age (Hauer 2019), HRSA HPSA designations, the Dartmouth Atlas ZIP–HSA–HRR crosswalk, and OpenStreetMap/Geofabrik road networks, each cited with dataset versions and retrieval dates in the Supplementary Material, which also describes the analytic steps and parameters. Individual physician-level records derived from these sources are not redistributed. Aggregate result tables (county- and state-level) are available from the corresponding author on reasonable request.

## Use of artificial intelligence

Study analyses (acquisition of the public source data, record linkage and classification auditing, geospatial simulation, statistical tabulation, and figure generation) were executed with the assistance of a generative artificial-intelligence system (Claude; Anthropic, San Francisco, CA) operating under the authors’ direction, specification, and review. All analyses were materialized as saved, rerunnable code with versioned inputs and outputs, and every reported value traces to those scripts (Supplementary Material S-M9).

## Attachments

Supplementary Material (supplementary methods S-M1–S-M10, Figures S1–S5, STROBE checklist, and data-source vintage register)

## REFERENCES

1. Rogers HW, Weinstock MA, Feldman SR, Coldiron BM. Incidence estimate of nonmelanoma skin cancer (keratinocyte carcinomas) in the U.S. population, 2012. JAMA Dermatol. 2015;151(10):1081–1086. PMID: 25928283.

2. Connolly SM, Baker DR, Coldiron BM, et al. AAD/ACMS/ASDSA/ASMS 2012 appropriate use criteria for Mohs micrographic surgery. J Am Acad Dermatol. 2012;67(4):531–550. PMID: 22959232.

3. National Comprehensive Cancer Network. NCCN Clinical Practice Guidelines in Oncology: Basal Cell Skin Cancer. Version 1.2026. Accessed September 12, 2026. https://www.nccn.org/guidelines/guidelines-detail?category=1&id=1416

4. National Comprehensive Cancer Network. NCCN Clinical Practice Guidelines in Oncology: Squamous Cell Skin Cancer. Version 2.2026. Accessed September 12, 2026. https://www.nccn.org/guidelines/guidelines-detail?category=1&id=1465

5. Steinman HK, Clever H, Dixon A. The characteristics of Mohs surgery performed by dermatologists who learned the procedure during residency training or through postgraduate courses and observational preceptorships. Proc (Bayl Univ Med Cent). 2016;29(2):119–123. PMID: 27034540.

6. Tierney EP, Hanke CW, Kimball AB. Recent changes in the workforce and practice of dermatologic surgery. Dermatol Surg. 2009;35(3):413–419. PMID: 19175662.

7. du Plessis PJ, Leventer M, Krekels G, de Wet JD, Laeuchli S. Outcomes of Mohs micrographic surgery at the American Society for Dermatologic Surgery International Traveling Mentorship Program International Mohs Fellowship Recognition units: a retrospective survey of 5889 cases from South Africa, Romania, and the Netherlands. Dermatol Surg. 2019;45(Suppl 2):S155–S162. PMID: 31764300.

8. Greif C, Burgess BA, Owen JL. Mohs micrographic surgery in the Veterans Health Administration: an inaugural nationwide assessment. Dermatol Surg. 2026 (epub). doi:10.1097/DSS.0000000000005262. PMID: 42430732.

9. Zhou AE, Gronbeck C, Sedghi T, Feng H. Comparative analysis of Mohs surgeon characteristics by societal membership, 2014 to 2022. Dermatol Surg. 2025;51(5):551–553. PMID: 39729097.

10. Gronbeck C, Sahin S, Feng H. Characteristics and geographic distribution of Mohs surgeons obtaining Micrographic Dermatologic Surgery board certification, 2021 to 2025. Dermatol Surg. 2026 (epub). doi:10.1097/DSS.0000000000005066. PMID: 41804989.

11. Centers for Medicare & Medicaid Services. Medicare Claims Processing Manual Transmittal 434 (CLIA edits for Mohs codes). January 14, 2005.

12. 42 CFR part 493 (Laboratory Requirements), §§493.1443-493.1449 (current text).

13. Riddle AO, Carucci JA, Criscito MC, Stevenson ML. Fellowship-trained Mohs surgeons as CLIA laboratory directors: navigating recent policy changes. Dermatol Surg. 2026 (epub). doi:10.1097/DSS.0000000000005043. PMID: 41650334.

14. Clinical Laboratory Improvement Amendments of 1988 (CLIA) Fees; Histocompatibility, Personnel, and Alternative Sanctions for Certificate of Waiver Laboratories; Final Rule. 88 Fed Reg 89976 (December 28, 2023).

15. Centers for Medicare & Medicaid Services. QSO-25-21-CLIA: CLIA enforcement discretion and clarification on personnel regulations. June 23, 2025.

16. Centers for Medicare & Medicaid Services; Centers for Disease Control and Prevention. Request for Information: Clinical Laboratory Improvement Amendments of 1988 (CLIA) Regulations. 91 Fed Reg 43586 (July 16, 2026).

17. Veterans Health Administration. Directive 1101.12: Mohs Micrographic Surgery. January 27, 2023.

18. Tam A, Yuan JT, Mauro TM, et al. Mohs micrographic surgery in the Veterans Health Administration. Fed Pract. 2018;35(Suppl 1):S38–S43. PMID: 30766388.

19. Feng H, Belkin D, Geronemus RG. Geographic distribution of U.S. Mohs micrographic surgery workforce. Dermatol Surg. 2019;45(1):160–163. PMID: 29620564.

20. Sharma AN, Peterman N, Juhasz M, Shive M. MMS hotspots: a cross-sectional comparison of U.S. counties with and without Mohs micrographic surgery. Arch Dermatol Res. 2023;316(1):21. PMID: 38060044.

21. Feng H, Berk-Krauss J, Feng PW, Stein JA. Comparison of dermatologist density between urban and rural counties in the United States. JAMA Dermatol. 2018;154(11):1265–1271. PMID: 30193349.

22. Sedghi T, Gronbeck C, Feng H. Comparison of socioeconomic and demographic characteristics of counties with and without Mohs micrographic surgeons: a cross-sectional Medicare analysis. Dermatol Surg. 2021;47(12):1655–1657.

23. Tsai S, Kooistra LJ, Conic RZ, Bordeaux JS. Trends in Medicare billing among Mohs surgeons in the United States during 2012: a retrospective cohort study. Dermatol Surg. 2019;45(2):268–273. PMID: 30199438.

24. Kodumudi V, Hales HA, Cohen JM, Feng H. Employment and migration patterns of recent Micrographic Surgery and Dermatologic Oncology fellowship graduates. Dermatol Surg. 2021;47(7):934–937. PMID: 33867465.

25. Davis B, Cleaver J, Meecham J, Taylor E. Bridging the gap: rural access to Mohs micrographic surgery and the role of training pathways. Dermatol Surg. 2026 (epub September 22, 2026). doi:10.1097/DSS.0000000000005360. PMID: 42766302.

26. von Elm E, Altman DG, Egger M, et al. The Strengthening the Reporting of Observational Studies in Epidemiology (STROBE) statement. Lancet. 2007;370(9596):1453–1457.

27. Centers for Medicare & Medicaid Services. Medicare Physician & Other Practitioners — by Provider and Service public use files, CY2020-CY2024 (dataset versions and retrieval dates in Supplementary Material).

28. Davis M, et al. Trends in the Mohs micrographic surgery workforce, 2013-2023. American Society for Dermatologic Surgery Annual Meeting; 2025 (conference report).

29. American Board of Dermatology. Newly certified Micrographic Dermatologic Surgery diplomates, 2021-2025 (public lists; archived captures and retrieval dates in Supplementary Material).

30. Perlis CS, Perlis RH. Accuracy of attestation among Micrographic Dermatologic Surgery diplomates. JAMA Netw Open. 2022;5(9):e2229795. PMID: 36053539.

31. US Department of Agriculture, Economic Research Service. Rural-Urban Continuum Codes, 2023.

32. 42 CFR 422.116 (Medicare Advantage network adequacy: time and distance standards), dermatology; CY2026 Health Service Delivery reference file.

33. Accreditation Council for Graduate Medical Education. ACGME Program Requirements for Graduate Medical Education in Micrographic Surgery and Dermatologic Oncology (specialty 081); Accreditation Data System public report, AY2026-2027.

34. San Francisco Match. Micrographic Surgery and Dermatologic Oncology Match: data report, 2015-2024. sfmatch.org (retrieved August 18, 2026).

35. American College of Mohs Surgery. MSDO match results and rank-list statistics, 2019-2025. mohscollege.org.

36. Centers for Medicare & Medicaid Services. 2011 Medicare Advantage network adequacy criteria development overview. Baltimore, MD: CMS; 2011. Accessed September 26, 2026. https://www.cms.gov/medicare/medicare-advantage/medicareadvantageapps/downloads/2011_ma_network_adequacy_criteria_overview.pdf

37. Centers for Medicare & Medicaid Services. Medicare Monthly Enrollment (annual county and state rows by age group, CY2013-CY2025). data.cms.gov; June 2026 data release, posted September 23, 2026 (retrieved September 25, 2026).

38. Hauer ME. Population projections for U.S. counties by age, sex, and race controlled to Shared Socioeconomic Pathway. Sci Data. 2019;6:190005. doi:10.1038/sdata.2019.5. PMID: 30720801. Data: Open Science Framework, doi:10.17605/OSF.IO/9YNFC (SSP2 county file, retrieved September 25, 2026).

39. Centers for Medicare & Medicaid Services. Medicare Physician & Other Practitioners — by Geography and Service public use files, CY2013-CY2024 (national rows, HCPCS 17311 and 17313). data.cms.gov (retrieved September 25, 2026).

40. Weldon Cooper Center for Public Service, Demographics Research Group. National population projections by age and sex, 2030-2050 (V2026, July 2026). University of Virginia; coopercenter.org (retrieved September 25, 2026).

41. Krishnan A, Xu T, Hutfless S, et al. Outlier practice patterns in Mohs micrographic surgery: defining the problem and a proposed solution. JAMA Dermatol. 2017;153(6):565–570. PMID: 28453605.

42. AMN Healthcare. 2025 Survey of Physician Appointment Wait Times and Medicare and Medicaid Acceptance Rates. Dallas, TX: AMN Healthcare; May 2025.

43. Lee J, Forrester VJ, Novicoff WM, Guffey DJ, Russell MA. Surgical delays of less than 1 year in Mohs surgery associated with tumor growth in moderately- and poorly-differentiated squamous cell carcinomas but not lower-grade squamous cell carcinomas or basal cell carcinomas: a retrospective analysis. J Am Acad Dermatol. 2022;86(1):131–139. PMID: 34499990.

44. Eide MJ, Weinstock MA, Dufresne RG, et al. Relationship of treatment delay with surgical defect size from keratinocyte carcinoma (basal cell carcinoma and squamous cell carcinoma of the skin). J Invest Dermatol. 2005;124(2):308–314. doi:10.1111/j.0022-202X.2004.23546.x. PMID: 15675948.

45. Sattler SS, Sharon VR. National trends in Mohs micrographic surgery: a decade of enhanced access, improved tumor clearance, and implications for future workforce planning. Dermatol Surg. 2026 (epub). doi:10.1097/DSS.0000000000005218. PMID: 42328957.

46. Alam M, Ibrahim O, Nodzenski M, et al. Adverse events associated with Mohs micrographic surgery: multicenter prospective cohort study of 20 821 cases at 23 centers. JAMA Dermatol. 2013;149(12):1378–1385. PMID: 24080866.

47. Mosterd K, Krekels GAM, Nieman FHM, et al. Surgical excision versus Mohs’ micrographic surgery for primary and recurrent basal-cell carcinoma of the face: a prospective randomised controlled trial with 5-years’ follow-up. Lancet Oncol. 2008;9(12):1149–1156. PMID: 19010733.

48. Grabski WJ, Salasche SJ, McCollough ML, et al. Interpretation of Mohs micrographic frozen sections: a peer review comparison study. J Am Acad Dermatol. 1989;20(4):670–674. PMID: 2715412.

49. Kesty K, Sangueza OP, Leshin B, Albertini JG. Mohs micrographic surgery and dermatopathology concordance: an analysis of 1421 Mohs cases over 17 years. J Am Acad Dermatol. 2023;88(1):118–122. PMID: 29246825.

50. Aggarwal P, Neltner SA, Fleischer AB Jr. Risk factors that are associated with outliers in Mohs micrographic surgery in the national Medicare population, 2018. Dermatol Surg. 2022;48(2):181–186. PMID: 34923533.

51. Albertini JG, Wang P, Fahim C, et al. Evaluation of a peer-to-peer data transparency intervention for Mohs micrographic surgery overuse. JAMA Dermatol. 2019;155(8):906–913. PMID: 31055597.

52. American Board of Dermatology. CLIA lab director qualifications amended to include ABD-certified dermatologists. abderm.org; 2025.

53. American Osteopathic Board of Dermatology; American Osteopathic College of Dermatology. Letter to CMS/CDC regarding CLIA laboratory director requirements. March 5, 2025.

54. American Academy of Dermatology Association. AADA advocacy win: CMS suspends CLIA lab director enforcement policy. June 23, 2025. Accessed September 12, 2026. https://www.aad.org/member/advocacy/update/2025/cms-suspends-clia-enforcement

55. American Society for Mohs Surgery. Message from ASMS President regarding changes in CLIA laboratory director qualifications. Member communication; April 2, 2025.

56. American Society for Dermatologic Surgery Association. New rules impacting high-complexity laboratories. ASDS/A Member News; April 4, 2025.

57. First Coast Service Options. Local Coverage Determination L33689: Mohs Micrographic Surgery (revision effective January 8, 2019).

58. Walocko FM, Memon R, Kwasny MJ. Micrographic dermatologic surgery (MDS) diplomates: a demographic evaluation and comparison of Medicare case volume. Arch Dermatol Res. 2022;314(2):213–216. PMID: 35133478.

59. Porras Fimbres DC, Rundle CW, Presley C, Stamey C. State-based population analysis of fellowship trained vs. non-fellowship trained Mohs surgeons. Arch Dermatol Res. 2023;315(4):1071–1073. PMID: 36484800.

