## Supplementary Material for "Access Consequences of Restricting Mohs Micrographic Surgery to Fellowship-Trained Surgeons: A National Workforce, Board-Certification, and Drive-Time Simulation Study"

This file contains: Supplementary Methods (S-M1–S-M10); Supplementary Figures S1–S5 (legends here; the figure pages follow at the end of this file); the STROBE checklist; and the data-source vintage register. Every number is generated by a saved, rerunnable script from the public sources listed in the data-source vintage register; analytic steps, parameters, seeds, and quality-control results are described in S-M1–S-M10.

---

### S-M1. Workforce census

Active Mohs surgeons were identified in the CMS Medicare Physician & Other Practitioners — by Provider and Service public use files (PUF), calendar years 2020–2024, as individual NPIs with ≥1 billed service of CPT 17311 or 17313 in a year; stage codes 17312/17314/17315 contributed volume and stages-per-case metrics. Organizational NPIs (entity code O; n=16) were excluded. Cases = services(17311)+services(17313). Two rosters were defined a priori: a **dynamics roster** (any billing 2020–2024; n=3,451 individuals) for attrition, cohort, and pipeline analyses, and a **simulation roster** (billing in 2023 and/or 2024; n=3,167) as the sole denominator for geographic and access simulation, so that pre-2023 exits do not overstate current capacity. CY2020 is COVID-distorted and was never used as a sole activity anchor. NPPES (the August 2026 full Data Dissemination file) supplied names, credentials, sex, and secondary practice locations; every distinct practice location observed in the window was retained and geocoded (Census batch geocoder, Nominatim fallback, manual resolution of institutional addresses; 100% resolved to coordinates + county + ZCTA; 97.6% street-level). **Suppression:** the PUF suppresses clinician-code-year rows under 11 beneficiaries. In 2024, 197 surgeons billed first-stage codes with no visible 17312 row (censored stage volumes), and left-tail extrapolation of the beneficiary distribution estimates ≈124 additional surgeons (≈4%) entirely invisible — plausibly low-volume, rural, and R/P-trained; this gap understates R/P-trained capacity and the reported access losses (the classification error described in S-M2 runs the opposite way).

**Self-designated specialty and MMS share of practice.** Specialty is the clinician's NPPES primary taxonomy (general dermatology 207N00000X; Mohs–Micrographic Surgery 207ND0101X; Procedural Dermatology 207NS0135X; Dermatopathology 207ND0900X). For each surgeon active in 2024, MMS stage services (CPT 17311–17315) were divided by the surgeon's total Medicare Part B medical services from the CY2024 Physician & Other Practitioners by-Provider summary file (all 3,042 active surgeons matched by NPI; CMS suppresses the medical-only subtotal for 964 surgeons, so medians use the unsuppressed subset — n = 1,032/938/30/78 for metro FT/metro R/P/nonmetro FT/nonmetro R/P — and a total-services sensitivity on all 3,042 was materially identical, 27.2/4.4/29.7/4.8%); same-day reconstructive repairs are excluded from the numerator, so the shares understate the Mohs-related fraction of practice in both pathways. Among nonmetropolitan R/P-trained surgeons, 110 of 155 (71.0%) self-designate as general dermatologists (57 of 114, 50.0%, among nonmetropolitan FT surgeons) and MMS is a median 5.3% (IQR 2.9–8.3) of Medicare services; FT surgeons overall, 27.9% (IQR 14.9–40.6).

### S-M2. Training-pathway classification and the adjusted (scenario B2) machinery

**Directory collections (all 2026-08-17):** the complete ACMS "Find a Mohs Surgeon" directory via (i) a 133-seed ZIP-radius grid (200-mile radius; coverage proof: every active US ZIP centroid within 190 miles of a seed; per-seed completeness audit with zero gaps) and (ii) a 1,728-query surname sweep of the directory's server-side substring search, which reaches members without listed addresses (358 such members found; 339 roster reclassifications). ASMS's public Surgeon Locator (46 states) provided an affiliation flag only; listing is opt-in, so ASMS affiliation is undercounted by design. ACGME's public program report (specialty 081) verified the fellowship-program universe (88 programs, AY2026-27).

**Matching:** tiered deterministic and fuzzy matching (exact name+state; nickname canon; accent/compound-surname handling; NPPES-suffix Jr/Sr resolution; manual adjudication of all sub-threshold candidates; 2,572 logged decisions). ACMS directory match rate 93.5%; unmatched directory entries explained (predominantly Kaiser/California, military, retirees, new members). External validation: an independent predecessor analysis using a different surname method counted 1,664 ACMS-listed 2024 billers vs our 1,665 (Δ=1).

**Audit and correction:** a seeded simple random sample of R/P-classified surgeons (n=30 after the surname sweep rebased the frame; seed 20260817) was web-audited with quoted evidence. Residual fellowship-trained share: 3/30 = 10.0% (Wilson 95% CI 3.46–25.62%); strict definition 2/30 = 6.7% (1.85–21.32%). This error is asymmetric — it can only move fellowship-trained surgeons into the R/P-classified group — so it enlarges the excluded group and overstates by-protocol exclusion impact, and the adjusted estimates below are correspondingly lower; the audit's public-bio method can miss fellowship training for surgeons with sparse web presence, so the audited rate is a lower bound on the true rate. **Scenario B2** propagates this error: the 3 audit-confirmed fellowship-trained surgeons are FT in every draw and the 27 confirmed R/P are R/P in every draw; every other R/P-classified surgeon reclassifies FT independently with probability p ∈ {0.1000 (point), 0.0346 (lower), 0.2562 (upper)}; 500 Monte Carlo draws per p (100 for the routing-intensive Phase-4 metrics) from a fixed-seed generator (numpy default_rng(20260817)), with a common-uniform-matrix design that nests the FT sets monotonically across p within each draw. The manuscript's main analyses use the by-protocol (directory-based) definition; adjusted (B2) results are consolidated in manuscript Table VI (Results) and carried throughout the supplementary tables (point [95% draw interval]; bounds = envelope at the Wilson limits). A volume-weighted variant (reclassification probability increasing in surgeon volume, marginal rate preserved) changed no conclusion.

### S-M3. Board-certification linkage

ABD public lists of newly certified MDS diplomates (2021: 1,677; 2022: 398; 2023: 357; 2024: 287; 2025: 299; 3,013 unique names after normalization/dedup) were harvested with archival provenance (Wayback Machine snapshots for 2021–2024; live capture 2025; byte-hash integrity checks). All 3,451 roster NPIs were then checked against ABD's public Dermatologist Search verification service, which accepts NPI queries — an authoritative census that replaced sampling inference (2,280 certified; 646 affirmatively not; 525 absent from the lookup). Name-matcher validation against the authoritative results: exact-match non-ambiguous precision 99.9% (2,223/2,225); lower tiers replaced by authoritative results; false-negative rate among non-matches 3.10%. Final flag: authoritative result where available; otherwise only exact non-ambiguous name matches counted as certified (232); everything else counted as uncertified. The 525-NPI gap stratum is 45% DO-credentialed (vs 10% roster-wide), consistent with

AOBD-certified osteopathic surgeons structurally absent from ABD's lookup, which understates certification, particularly among R/P-trained surgeons. All certification shares are minimums.

### S-M4. Geography

County reference: 2023 county-equivalents (Connecticut planning regions; 52 CT site rows reassigned by point-in-polygon of existing coordinates). RUCC 2023; county population = Census Vintage-2025 estimates; beneficiaries = CMS Medicare Monthly Enrollment CY2025 annual county averages; median household income = SAIPE 2022 (via the USDA ERS county file); primary-care HPSA area designations = HRSA snapshot 2026-08-17. Puerto Rico's 7 surgeons were tabulated separately and excluded from county-class/density/national-simulation universes. Surgeons attribute to **all** practice sites (primary-site sensitivity retained throughout; volumes split equally across sites in geographic sums). County access classes: R/P-only, FT-only, mixed. Market robustness: Dartmouth Atlas ZipHsaHrr19 crosswalk (site-ZIP coverage 99.80%); 2020 Census ZCTA populations; market beneficiaries apportioned by within-county ZCTA population shares. The market view is more severe than the county view: only 12% of R/P-only-county residents belong to a hospital market containing any by-protocol FT surgeon, and 197 of the 223 R/P-only counties sit wholly within R/P-only Hospital Service Areas. Note the population-vintage difference (county tables: Vintage-2025 estimates; market tables: 2020 Census) wherever both appear.

### S-M5. Drive-time simulation

**Origins:** 26,419 populated-ZCTA population-weighted centroids (2020 Census Centers of Population block-group means, point-in-polygon to 2020 ZCTA polygons), carrying 331.4M residents and 68.1M county-allocated beneficiaries. **Routing:** OSRM v6.0.0 (car profile, MLD) over Geofabrik OpenStreetMap extracts dated 2026-08-16, as 31 overlapping regional graphs; main-road network (residential/service ways dropped) — a measured deviation: two regions rebuilt on the full drivable network re-routed identical pairs with mean bias under 1 minute (90% of pairs within ±5 minutes), second-order against the +6.8-minute headline effect. **Validation:** 10 origin–destination pairs against public mapping estimates — route distances agree; times deviate mean |12| minutes (metro slightly fast, rural 13–24% slow: free-flow speed model, no congestion), symmetric across scenarios, so scenario contrasts are insulated. **Candidate cache:** each origin routed to its 25 nearest sites overall plus 15 nearest by-protocol-FT sites (great-circle prefilter; AK/HI/PR restricted in-state); because every scenario's surviving set contains the by-protocol-FT set, the nearest surviving site under any scenario/draw lies inside the cache (invariant asserted over 1,000 origins × 100 draws; zero violations). **No-road-access stratum:** strict 25-km snap radius, per-coordinate failure parsing, a physical-validity guard (road distance ≥ great-circle), and a curated ferry-dependent-island rule prevent fabricated routes; 105 origins (466,467 people) have no drivable path to any Mohs site at baseline and are excluded from mean/median statistics, counted beyond every threshold, and disclosed on every figure. 154,100 Hawai'i residents (islands whose only in-island sites are R/P-staffed) lose all road-accessible access under scenario B — a categorical loss, not a longer drive. **Network adequacy:** 42 CFR 422.116 dermatology time-and-distance maxima (Large Metro 20 min/10 mi; Metro 45/30; Micro 60/45; Rural 75/60; CEAC 110/100; eCFR text current as of 2026-08-13) applied via each county's CY2026 Health Service Delivery designation; compliance = any surviving site within both maxima. We report populations living beyond the standards' maxima (the regulation's own 85%-of-beneficiaries test applies plan-by-plan). **Scenario B3 (absorption):** surviving-FT spare capacity = headroom × own 2024 volume (+20% base; 0%/+50% sensitivity); excluded volume assigned by global greedy over cached site-to-site drive times.

### S-M6. Pipeline and replacement model

Pipeline supply from SF Match official data (2015–2024), ACMS match publications (2019–2025; row-count method reproduces SF Match 2024 exactly), and the ACGME program report. The 88 accredited MSDO programs are geographically concentrated (CA 9, PA 8, TX 7, NY 7, OH 6, MA 5, FL 5); 17 states, DC, and every territory have none; 32%/53% of graduates locate within 10/100 miles of their fellowship site (manuscript ref. Kodumudi et al.). Graduation years from the CMS Doctors and Clinicians National Downloadable File (92.9% linkage; sex concordance 99.8%; differential missingness among exited surgeons documented — current-roster age statistics minimally affected). Age = year − graduation year + 26. Replacement model: one two-compartment cohort model (pre-65 / 65+, with the turning-65 flow from graduation years) applied identically to no restriction, immediate exclusion, and grandfathering. Attrition, primary analysis: exit hazards measured from the 2021–2023 panel (rural FT 0.58%/yr before 65 and 9.4%/yr after; rural R/P 4.77%/yr and 10.5%/yr) — the calibration that reproduces observed net workforce growth and the observed share of active surgeons aged 65+; sensitivity analysis: retirement at a fixed age (65; 62/68), with surgeons already 65+ in 2026 leaving over ≈2 years. Rural FT arrivals = graduates × p(nonmetro), joining the pre-65 stock and subject to the same attrition as incumbents; R/P arrivals (no restriction only) at the observed 2021–2024 rate (34 over 2021–2024; 8.5 per year); p(nonmetro) = 10.3% (observed 2021–2024 FT entrants, 37/359; Wilson 95% CI 7.6–13.9%); pessimistic 6.7% = the long-run FT stock share (114 surgeons with a nonmetropolitan site among all FT surgeons on the simulation roster), the rate entrants revert to if mid-career migration toward metropolitan practice continues; optimistic 12% = the rising graduation-cohort trend continued one step past the newest cohort (nonmetropolitan share of FT surgeons by medical-school graduation cohort: ≤1989 2.6% [4/155]; 1990–99 4.0% [12/302]; 2000–09 6.2% [33/533]; 2010–14 8.0% [28/349]; 2015–19 10.6% [34/320]; by NPI-enumeration cohort, 2018+ 11.4% [13/114]) — a parameter-uncertainty bracket spanning what graduates have actually done, not a policy target; arrival volumes at the mature rural means (FT 226; R/P 190); demand frozen at 2024 (favors replacement). A threshold analysis (measured hazards, base pipeline of 95 graduates/yr; heads) sweeps p(nonmetro) to find the rate at which the 2026 rural headcount is restored within 5, 7, 10, and 15 years: 34.0%, 25.0%, 18.0%, and 13.0% of graduates, respectively, against an observed 10.3%. Scripts 53_harmonized_projection.py (engine; outputs harmonized_projection.csv, phase5_harmonized.json) and 54_county_survival_harmonized.py (phase5_county_curves_harmonized.json). **Grandfathering (scenario G):** R/P entry set to zero; existing R/P surgeons retained with exit hazards measured from the 2021–2023 panel (FT 1.18%/yr; R/P 6.70%/yr) in a two-compartment (pre-65 / 65+) calibration that reproduces observed net workforce growth; county-level loss uses per-surgeon ages and an incumbent-based replacement channel measured without definitional circularity (2021–2024 arrivals into counties R/P-only on pre-2021 incumbents: FT 0, R/P 39; one-sided 95% upper bound for the FT rate 0.0042/county-year).

### S-M7. Statistical reporting

Descriptive statistics with exact denominators; Wilson 95% CIs for audited proportions; Monte Carlo 95% draw intervals for adjusted estimates; standardized mean differences for county comparisons; surgeon-level logistic regression of pathway on nonmetropolitan practice with state fixed effects (primary-site attribution). No hypothesis tests are reported where the estimand is a census quantity. Analyses in Python (pandas/numpy); fixed seeds throughout.

### S-M8. Ethics

The study used exclusively public administrative, directory, and certification data about physicians in their professional capacities; no patient-level data were accessed.

---

### S-M9. Use of artificial intelligence

**Tool.** Claude (Anthropic, San Francisco, CA; the models used across the sessions were Claude Opus 4.6, Claude Opus 4.8, Claude Opus 5, Claude Fable 5, and Claude Fable 5.1, accessed through Anthropic's Claude desktop application and Claude Code) was used by the authors as an automation tool in research sessions between June and September 2026. Session-level records are retained by the authors; the resulting parameters, vintages, seeds, and quality-control results are recorded in this Supplement and in the study's parameter log.

**What the authors did.** The authors designed the study and specified each phase of the analysis in writing: the census window and inclusion rules, the training-pathway definition and its audit protocol, the geographic classifications and the access standards applied, the exclusion, capacity, and replacement scenarios, and the sensitivity analyses. They reviewed the output of every phase, directed the revisions, made every interpretive judgment, and are responsible for the argument of the paper.

**What the tool did.** Working to those specifications, the AI system automated the mechanical work of the analysis — the work a programmer or analyst would otherwise have carried out to the same instructions: writing and executing the analysis code; retrieving the public source data from portals and APIs (CMS PUF and by-Provider files, NPPES, ABD diplomate lists, ACMS/ASMS directories, Census/USDA/HRSA/Dartmouth references — vintages in the register below); running the record linkage, tiered matching, and audit machinery for pathway classification (S-M2); running the OSRM road-network drive-time simulation, the county/market access tabulations, the Monte Carlo correction propagation (fixed seeds, recorded parameters), and the capacity/pipeline models (S-M4-S-M6) and the demand projection (S-M10); tabulating results; and rendering the figures from the committed datasets. Every reported value derives from saved, rerunnable scripts operating on versioned raw archives (checksummed where retrieved in-session); the figures are data visualizations produced computationally from those datasets, and no generative-AI imagery was created or used.

**Role in writing (ICMJE: acknowledgment/declaration).** Manuscript text was drafted with AI assistance from the authors' outline, directives, and the study's tabulated outputs, then critically reviewed, edited, and verified by the authors, who take full responsibility for the content. All 58 references were verified against primary sources (held full texts or publisher/PubMed listings); no reference was generated without verification.

**Accountability.** No AI system is an author; the authors accept full responsibility for the integrity, accuracy, and originality of the work, consistent with ICMJE recommendations and the publisher's AI policies. The corresponding declaration appears on the title page.

### S-M10. Need for rural Mohs capacity and the demand projection (Table VII; Figure 7; version 2)

**Question and definition.** The replacement model of S-M6 holds demand at its 2024 level and treats today's rural access as the target. S-M10 estimates need in the sense used in health workforce planning, the rural Mohs capacity that keeps status quo access, meaning rural in-county Mohs surgery at today's rate per beneficiary as rural Medicare beneficiaries age and Mohs use per beneficiary grows, and brings rural residents inside the access standard used throughout the study, the 42 CFR 422.116 dermatology time-and-distance standard (described under "The standard" below). Rural counties are the nonmetropolitan county-equivalents (RUCC 4–9) of the 50 states and DC. Need in year t is $D(t) = (V + G) \times \Sigma_a w_a \times B_a(t)/B_a(2026) \times (1 + g)^{(t - 2026)} \times A$, where V = 53,232 is status quo access in 2026, the rural in-county first-stage volume of the cohort model (the 2024 census volume of nonmetropolitan sites, the engine's 2026 base); G = 63,836 is the access gap defined below; $B_a(t)$ is the projected number of rural fee-for-service beneficiaries in age band a (under 65, 65–74, 75–84, 85 and over); $w_a$ is the band's share of rural Mohs cases (0.022, 0.372, 0.436, 0.170: the age-specific rates $r_a$ below × rural beneficiaries by band, normalized); g is the growth in first-stage cases per fee-for-service beneficiary at fixed age; and A is an appropriateness factor (1.0 primary; 0.95 and 0.90 as sensitivities). Status quo access alone (G = 0), the version-1 target, is carried as its own row of Table VII and as the thin lines of Figure 7. Rural Mohs surgeons needed $R(t) = D(t) \div 197.9$, the 2024 mean caseload of surgeons at nonmetropolitan sites (53,232 ÷ 269), held fixed like the engine's per-surgeon volumes; because a site placed to serve a sparsely settled area rarely reaches that caseload, the surgeon figures are lower bounds on the sites required. Surgeons and cases are Mohs surgeons and first-stage Mohs cases (CPT 17311 + 17313) throughout; stages per case do not enter, so the post-2016 decline in stages per case is irrelevant here. Need so defined is the capacity that would have to exist within reach for rural residents to have the access the standard defines. It is not clinical need, and the residents it counts may obtain care today by traveling farther than the standard allows; at the national age-specific rates, rural fee-for-service beneficiaries as a whole would generate about 271,000 first-stage cases in 2026, about five times what rural counties deliver, because most rural residents are treated in metropolitan counties.

**The standard.** 42 CFR 422.116 sets, for each of five county types defined by population size and density, the maximum travel time and distance to the nearest network provider of each specialty that a Medicare Advantage plan must meet for 90% of a county's beneficiaries (85% in micropolitan, rural, and CEAC counties). For dermatology the maxima are 20 minutes and 10 miles (large metro), 45 and 30 (metro), 60 and 45 (micropolitan), 75 and 60 (rural), and 110 and 100 (counties with extreme access considerations), and a resident is inside the standard when a site lies within both limbs. CMS built the criteria for the contract year 2012 applications by mapping beneficiary locations against provider practice locations and testing the share of beneficiaries with a provider of each specialty within candidate travel times and distances in each county type (CMS, 2011 Medicare Advantage network adequacy criteria development overview, cited in the manuscript's Methods), so the values describe the prevailing pattern of care, the travel within which most beneficiaries could already reach a provider, rather than a clinical threshold; they were codified by rulemaking in 2020 (85 FR 33796), when the non-urban share was lowered from 90% to 85% and a 10-percentage-point telehealth credit was added for dermatology among other specialties. Three consequences for the estimate follow. The standard already concedes longer travel where population is sparse, so a nonmetropolitan resident counted outside it is outside a limit set for that geography. It counts a site within reach as access regardless of the site's capacity or waiting time, so need built on it is a lower bound in that respect. And no Mohs-specific standard exists; dermatology is the specialty under which Mohs surgery is delivered and billed, and the two-hour row of Table VII bounds the objection that patients reasonably travel farther for the procedure.

**The access gap (G).** Residents outside the standard today are those of Phase 4 scenario A (S-M5): 23,701,434 people and 5,367,661 beneficiaries in the routable origins outside Puerto Rico, of whom 12,204,065 residents and 3,128,083 beneficiaries live in nonmetropolitan counties (975 of the 1,958) and 11,497,369 residents and 2,239,578 beneficiaries in metropolitan counties. For each nonmetropolitan county, the share of its beneficiaries living in origins outside the standard (origin beneficiaries are county totals allocated by ZCTA population) multiplies the county's CY2025 fee-for-service beneficiaries by age band (below), and the national age-specific first-stage rates $r_a$ (below) convert the result to cases: $G = \Sigma_c s_c \times \Sigma_a FFS_a(c) \times r_a$ = 63,836 first-stage cases per year, or 323 rural Mohs surgeons at the 2024 rural mean caseload, against 269 surgeons practicing at nonmetropolitan sites today. The same calculation over all counties gives 101,608 cases (513 surgeons); the metropolitan part is outside the rural frame of Figure 7 and Table VII. The gap is grown with the same aging ratios and per-beneficiary growth as the in-county volume. As a sensitivity on the standard itself, the gap was recomputed counting only origins whose nearest Mohs surgeon is more than 120 minutes away by road (drive time only; the distance limb of the standard set aside): 2,467,026 residents nationally, of whom 2,078,808 residents and 505,473 beneficiaries live in 308 nonmetropolitan counties, generating 12,548 first-stage cases per year (63 surgeons). That two-hour gap is carried as a row of Table VII. Doubling each county type's own 422.116 time limit instead (120, 150, and 220 minutes for micropolitan, rural, and CEAC counties) leaves 711,735 nonmetropolitan residents outside and a gap of 3,721 cases (19 surgeons); it is recorded in access_gap_2026.json and not tabulated.

**Denominators.** The Physician & Other Practitioners files count only fee-for-service Part B beneficiaries, so the rates and the trends are per fee-for-service Part B beneficiary (Medicare Monthly Enrollment field B_ORGNL_MDCR_BENES) and need is in fee-for-service case units, the units of the cohort model's capacity. The enrollment file does not split age bands by coverage type, so each county's 2024 fee-for-service share was applied to every band and held constant; an assumption about Mohs use among Medicare Advantage enrollees therefore enters numerator and denominator alike and cancels out of $R(t)$ and the entry required. The fee-for-service share of beneficiaries fell from 64% to 42% over 2013–2024 (50 states and DC) and was 48.6% in nonmetropolitan and 40.4% in metropolitan counties in 2024.

**Enrollment by age ($B_a(2025)$).** CMS Medicare Monthly Enrollment, annual county rows CY2013–CY2025 from the June 2026 data release (posted 2026-09-23; retrieved 2026-09-25), with the dataset's ten age bands collapsed to four (under 65 = under 25 + 25–44 + 45–64; 65–74 = 65–69 + 70–74; 75–84 = 75–79 + 80–84; 85 and over = 85–89 + 90–94 + 95 and over). Age cells suppressed for small counts (3,427 county-band cells in 2025) take an equal share of the residual between the county total and its unsuppressed bands; 15 counties without a usable 2024 fee-for-service share take their state's share. Rural fee-for-service beneficiaries in 2025: 0.668 M under 65, 2.769 M aged 65–74, 1.682 M aged 75–84, and 0.543 M aged 85 and over (5.54 M of 11.64 M rural beneficiaries).

**Aging ($B_a(t) / B_a(2026)$).** Hauer's county projections by age, sex, and race controlled to Shared Socioeconomic Pathway 2 (manuscript ref. Hauer 2019; Open Science Framework project 9ynfc, file SSP_asrc.zip, retrieved 2026-09-25), summed over sex and race to the four bands (18 five-year age groups; 85 and over is the top group) for every projection year 2020–2045 and interpolated geometrically between five-year points. Growth ratios, never projection levels, are applied to the CY2025 enrollment, so the Medicare-versus-population level mismatch does not enter. Counties absent from the projections (seven Alaska census areas and two Connecticut planning regions; the projections use Connecticut's legacy counties) take their state's ratio; the under-65 band (largely beneficiaries entitled by disability) is held at its 2025 level. Rural growth 2026 → 2041: 65–74 × 0.80, 75–84 × 1.18, 85 and over × 1.48. Sensitivity: Weldon Cooper Center state projections by age and sex (V2026, July 2026; 2030/2040/2050 tables, interpolated), giving × 0.96, × 1.36, × 1.59. Cross-check recorded at acquisition: the projections' national 65-and-over totals are 0.5% (2025) and 4.95% (2040) above the Census Bureau's 2023 national projections (middle series), inside the ±5% tolerance fixed before the comparison; nothing was adjusted.

**Age-specific rates (r_a).** The prespecified ecological regression of county first-stage cases on beneficiaries by age band (no intercept; Poisson with identity link and non-negative least squares; metropolitan counties with at least two surgeons, all metropolitan counties, all counties, and a state panel for 2022–2024) was not identified: band shares are nearly collinear across geographies and cases follow the county of the practice rather than of the patient, so the fitted rates flipped between zero and implausible values across specifications (whole bands driven to zero while a neighbouring band absorbed them; the fits are retained as a diagnostic in demand_components.json). The age gradient was therefore taken from the Medicare patients of Mohs-dominant surgeons. The CY2024 Physician & Other Practitioners by-Provider file's beneficiary age counts (Bene_Age_LT_65/65_74/75_84/GT_84_Cnt, all services) for all 12,999 clinicians with a dermatology or micrographic-surgery specialty were joined to the roster by NPI (3,003 of the 2024 surgeons matched); among the 913 surgeons whose first-stage Mohs beneficiaries were at least 70% of all their Medicare beneficiaries (47% of national first-stage cases), the patient age shares were 1.9% under 65, 36.9% aged 65–74, 43.2% aged 75–84, and 18.0% aged 85 and over (thresholds of 50% and 90%: 2.2/37.8/42.8/17.3% and 1.6/36.7/43.6/18.2%). CMS suppresses an age cell under 11 beneficiaries and counter-suppresses a second, for Mohs surgeons usually the 85-and-over cell alongside the under-65 cell; each surgeon's suppressed remainder (total beneficiaries minus the visible cells) was spread over the missing cells in the pooled proportions of complete rows (complete rows alone give 2.0/37.2/42.4/18.5%). Cases per patient were taken as equal across bands; they rise with age, so the gradient is if anything understated. Applying the shares to the 2024 national first-stage total (1,140,695 cases) over national fee-for-service beneficiaries by band gives r_a = 7.4, 30.6, 59.0, and 71.3 cases per 1,000 fee-for-service beneficiaries under 65, 65–74, 75–84, and 85 and over (ratios 75–84 / 65–74 = 1.93; 85 and over / 65–74 = 2.33; the national check reproduces 1,140,695 exactly). The rates set the weights w_a and price the access gap; the in-county volume is anchored to observed rural supply.

**Growth in cases per beneficiary (g).** The primary rate is external and deliberately restrained: the age-adjusted rate of all skin-cancer procedures per fee-for-service beneficiary rose from 6,075 to 7,320 per 100,000 between 2006 and 2012 (Rogers et al., JAMA Dermatol 2015;151:1081–1086, manuscript ref. 1), 3.16% per year, and using it assumes that Mohs use grows only with the treatment of keratinocyte carcinoma and not through any further rise in the Mohs share of that treatment. The study's own data show faster growth. National first-stage services (CPT 17311 + 17313, facility and non-facility rows) from the Physician & Other Practitioners by Geography and Service files, CY2013–CY2024 (retrieved 2026-09-25), over national fee-for-service Part B beneficiaries: the standardized incidence ratio (each year's observed cases over the cases expected at the 2024 age-specific rates and that year's band structure) rose from 0.546 in 2013 to 1.0 in 2024, and the regression of log(ratio) on year with indicator terms for 2020 and 2021 gives +5.58% per year (standard error 0.12%; 2020 −10.7%, 2021 −3.5%); the crude rate rose from 21.3 to 40.8 first-stage cases per 1,000 fee-for-service beneficiaries. Part of that trend is selection: as Medicare Advantage drew beneficiaries out of fee-for-service, the population remaining in fee-for-service changed in ways age standardization does not remove. A state panel (50 states and DC, 612 state-years, 2013–2024, state fixed effects, weighted by fee-for-service beneficiaries) separates the two: $\log(SIR) = a_s + g \cdot t + \beta \cdot \log(\text{fee-for-service share})$ + COVID indicators gives $\beta = -0.32$ (SE 0.07), so a 10% fall in a state's fee-for-service share raises its fee-for-service Mohs rate by about 3.3%, and g = +4.42% per year (SE 0.30) with the share held constant, against +5.76% without the share term; the same fit on the all-beneficiary denominator returns the identical 4.42% ($\beta = +0.68$), as the algebra requires, and the pre-2020 window alone shows no detectable share effect. On an all-beneficiary denominator the simple trend is +1.45% per year; that series is diluted rather than bounded, because beneficiaries who move to Medicare Advantage leave the numerator while remaining in the denominator, and Medicare Advantage members use Mohs surgery at about the fee-for-service rate (24.5 vs 24.8 per 1,000 in 2014: Rogers HW, Beveridge M, Puente J, et al. J Am Acad Dermatol 2021;85:741–743). Table VII therefore carries the ladder: aging alone, 1.45%, the primary 3.16%, 4.42%, and 5.58%.

**Appropriateness (A).** A = 0.95 and 0.90 reduce every case count by 5% and 10%, an assumption of the order of the reduction achieved among outlier surgeons by the specialty's physician-level data-transparency intervention (manuscript ref. Albertini et al. 2019), applied here to all cases as a deliberately harsh assumption. Under A = 0.90 the entry required is 67 per year, more than three times observed entry.

**Entry required.** The cohort engine of S-M6 (script 53) was replicated and gated against harmonized_projection.csv (both attrition modes; exact). Because need exceeds rural supply from 2026 on, the entry required is the constant annual rural entry that brings rural volume up to need by 2041 (the every-year criterion is infeasible at t = 0); for status quo access alone the two criteria coincide. For both pathways together, fellowship arrivals were held at the observed 9.785 per year (95 graduates × 10.3%) and the R/P rural entry x found by bisection; the entry required is 9.785 + x. For the fellowship pipeline alone with the R/P workforce retained (the structure of a restriction with grandfathering, R/P entry closed), graduates per year at 10.3% rural siting were found by bisection and also expressed as the rural siting rate at 95 graduates per year; with the R/P workforce removed (the exclusion structure) likewise. Arrivals practice at the mature rural means (fellowship-trained 226 cases, R/P-trained 190) with pathway-specific exit hazards (S-M6). Observed entry, 2021–2024: 37 fellowship-trained and 34 R/P-trained rural entrants, about 18 per year. Cumulative shortfall is $\Sigma_t \max(0, D(t) - \text{volume}(t))$ under measured exit hazards. Results by row are in manuscript Table VII; against status quo access alone the pipeline as it stands keeps pace only while g stays below 2.27% per year, a restriction with grandfathering only below 0.60%, and immediate exclusion at no rate, because its 2041 supply (53,292 cases) is below the aging-only need (57,629). With the gap counted only beyond two hours, the entry required is 34.2 per year (24.4 R/P-trained), the fellowship pipeline alone 245 graduates per year (27% rural siting) with the R/P workforce retained and 275 (30%) with it removed, and the cumulative shortfall 353,000, 509,000, and 794,000 cases under no restriction, a restriction with grandfathering, and immediate exclusion. The caseload sensitivity multiplies every surgeon's volume by $1.02^{(t - 2026)}$ (the national median caseload rose from 260 in 2021 to 285 in 2024). The cumulative shortfall against demand held at its 2024 level (the difference form of Figure 5) is 441,000 fewer rural cases under immediate exclusion (pipeline band 351,000–573,000) and 157,000 under a restriction with grandfathering than with no restriction (Figure S5).

**Network-adequacy trajectory (Figure 7, C).** Residents living outside the 42 CFR 422.116 dermatology time-and-distance standard were projected by year and scenario from the S-M5 drive-time matrix (26,188 routable origins outside Puerto Rico, 330.9 M residents, each origin's 40 cached candidate sites) with no new routing. Under no restriction and a restriction with grandfathering, the practice sites of an R/P-only county go dark in the year the county loses its last surgeon in the county-survival draws of S-M6 (500 draws; a county rescued by an arrival before its last exit keeps its sites), and every other site is held active. Under immediate exclusion the lower edge holds the scenario-B site set fixed (46.8 M residents) and the upper edge also darkens the 96 fellowship-served nonmetropolitan counties (114 fellowship-trained surgeons) at the measured fellowship-trained exit hazards with no replacement. The 2026 values reproduce S-M5 exactly (23,701,434 and 46,787,946 residents; 5,367,661 beneficiaries under no restriction). Because only those counties change, the trajectories understate the decline in adequacy. Results: no restriction 28.2 M [26.8–30.2] residents outside the standard by 2041, a restriction with grandfathering 30.9 M [29.2–33.0], immediate exclusion 46.8–47.6 M; fixed-age retirement 26.2, 27.3, and 46.8–48.2 M.

**Scripts and outputs.** 57_figure7_deficit.py (scenarios relative to no restriction at 2024 demand; Figure S5), 58_adequacy_trajectory.py (Figure 7, C), 59_demand_components.py (rates, aging, trends), 66_trend_share_adjusted.py (the state-panel share adjustment and the breakeven rates), 67_access_gap_requirement.py (the access gap under the standard and beyond two hours, and the need series), 61_table7_requirement.py (Table VII), 62_figure7_final.py (Figure 7, Figures S4–S5), and 68_figure5_relabel.py (Figure 5 with the scenario name written "restriction with grandfathering"); outputs demand_components.json, trend_share_adjusted.json, access_gap_2026.json, rural_requirement_projection.csv, adequacy_trajectory.csv, table7_summary.json, and fig7_final_summary.json. Every reported figure is re-derived from these committed files.

### Supplementary Figure Legends

**Figure S1.** Drive time to the nearest Mohs surgeon, status quo (scenario A). Road-network minutes from populated-ZCTA population-weighted centroids; no-road-access origins hatched.

**Figure S2.** Drive time to the nearest Mohs surgeon under by-protocol exclusion (scenario B).

**Figure S3.** Population-weighted survival curves of drive time under scenarios A, B, and B2, with 30/60/90/120-minute thresholds and the no-road-access stratum reported separately.

**Figure S4.** Rural Mohs capacity in levels against need, 2026–2041 (the level form of manuscript Figure 7, A–B). A, Rural Mohs surgeons, and B, annual rural first-stage Mohs cases, by scenario (cohort model of Figure 5) against two grey lines, both at the 2006–2012 growth of skin-cancer treatment and county aging. The thick grey line is need as defined in Figure 7, status quo access kept and the nonmetropolitan beneficiaries outside the CMS dermatology standard brought inside it; the thin grey line at the lower edge of the shading is status quo access alone, rural in-county Mohs surgery kept at today's rate per beneficiary with nothing added for residents already outside the standard. Need is 592 surgeons and 117,100 cases in 2026 and 1,021 surgeons and 202,000 cases in 2041 (status quo access alone, 464 and 91,800); in 2041 no restriction reaches 386 surgeons and 80,600 cases per year, a restriction with grandfathering 294 and 63,000, immediate exclusion 236 and 53,300 (adjusted classification 248 and 55,100). C, as manuscript Figure 7, C. Each scenario is drawn twice in its own color, as a solid line in which surgeons exit at the measured hazards (primary analysis) and as a dotted line in which they retire at a fixed age of 65 (sensitivity analysis); the dashed green line is immediate exclusion under the adjusted classification; the band spans pessimistic to optimistic pipeline assumptions in A and B and the Monte Carlo 95% interval in C.

**Figure S5.** The restriction scenarios relative to no restriction with demand held at its 2024 level, 2026–2041 (the difference form of Figure 5). Each curve is the no-restriction trajectory minus the scenario trajectory. A, Rural Mohs surgeons, and B, rural first-stage Mohs cases per year: immediate exclusion starts at the excluded stock (155 surgeons; 27,700 cases) and a restriction with grandfathering grows from zero as R/P surgeons retire unreplaced — 150 surgeons and 27,300 cases per year, and 93 and 17,600, in 2041 (adjusted classification, exclusion: 144 and 26,400); summed over 2026–2041, 441,000 (pipeline band 351,000–573,000) and 157,000 fewer rural cases than with no restriction. C, Residents of counties without an in-county Mohs surgeon beyond no restriction: all 223 R/P-only counties (23.4 million) under immediate exclusion at once, narrowing to 16.2–16.9 million by 2041 because no restriction itself loses 77 counties; a restriction with grandfathering 3.5 [2.3–4.9] million by 2041 (adjusted classification 3.1 million). Each scenario is drawn twice in its own color, as a solid line in which surgeons exit at the measured hazards (primary analysis) and as a dotted line in which they retire at a fixed age of 65 (sensitivity analysis); the dashed green line is the adjusted classification (immediate exclusion in A and B, grandfathering in C); the band spans pessimistic to optimistic pipeline assumptions in A and B and the Monte Carlo 95% interval for grandfathering in C.

---

### STROBE checklist (cross-sectional)

| Item | # | Where addressed |
|---|---|---|
| Title/abstract | 1 | Title page; structured abstract (design named; robustness statement) |
| Background/rationale | 2 | Introduction ¶1–2 |
| Objectives | 3 | Introduction ¶3 |
| Study design | 4 | Methods (census + cross-sectional linkage + simulation) |
| Setting | 5 | Methods (US Medicare, 2020–2024; vintages in S-M1/S-M4 and vintage register) |
| Participants | 6 | Methods: eligibility = ≥1 billed 17311/17313; dual-roster definitions (S-M1) |
| Variables | 7 | Methods + S-M2–S-M6, S-M10 (pathway, certification, rurality, drive time, adequacy, projected demand) |
| Data sources/measurement | 8 | S-M1–S-M6, S-M10; vintage register |
| Bias | 9 | Audited misclassification with Monte Carlo propagation (S-M2); suppression, FFS lens (S-M1; Limitations) |
| Study size | 10 | Census (complete enumeration); audit n justified by frame rebase (S-M2) |
| Quantitative variables | 11 | Methods; S-M7 |
| Statistical methods | 12 | S-M7 |
| Participants (results) | 13 | Results ¶1 (3,042/3,167/3,451 with definitions) |
| Descriptive data | 14 | Tables I–II |
| Outcome data | 15 | Tables II–IV, VII |
| Main results | 16 | Results (directory-based definition); adjusted re-derivation in Results (Table VI) |

| Item | # | Where addressed |
|---|---|---|
| Other analyses | 17 | HSA/HRR robustness; volume-weighted variant; primary-site sensitivity; scenarios B3/G; demand-projection sensitivities (Table VII) |
| Key results | 18 | Discussion ¶1 |
| Limitations | 19 | Discussion, Limitations |
| Interpretation | 20 | Discussion |
| Generalizability | 21 | Limitations (Medicare FFS lens) |
| Funding | 22 | Title page |

---

### Data-source vintage register

| Source | Vintage / version | Role |
|---|---|---|
| CMS Medicare Physician & Other Practitioners PUF | CY2020–CY2024 (catalog modified 2026-05-21) | Census, volumes |
| CMS Medicare Physician & Other Practitioners — by Provider | CY2024 (UUID 8889d81e-2ee7-448f-8713-f071038289b5; retrieved 2026-08-18) | Practice-mix denominators |
| NPPES Data Dissemination | V.2 August 2026 | Names, credentials, secondary locations |
| ACMS Find-a-Surgeon directory | Harvested 2026-08-17 (ZIP-grid + surname sweep) | FT classification |
| ASMS Surgeon Locator | Harvested 2026-08-17 | Affiliation flag |
| ABD MDS diplomate lists + Dermatologist Search | Cohorts 2021–2025; NPI sweep 2026-08-18 | Certification |
| USDA ERS RUCC | 2023 | Rurality |
| Census county population estimates | Vintage 2025 | Denominators |
| CMS Medicare Monthly Enrollment | CY2025 annual county averages (catalog 2026-07-23) | Beneficiaries |
| SAIPE median household income (via ERS) | 2022 | County covariate |
| HRSA primary-care HPSA designations | Snapshot 2026-08-17 | County covariate |
| Census Centers of Population; 2020 ZCTA polygons | 2020 | Origins |
| Geofabrik OSM extracts / OSRM v6.0.0 | 2026-08-16 | Routing |
| 42 CFR 422.116 + CY2026 HSD reference file | eCFR 2026-08-13; HSD 2025-12-17 | Network-adequacy standards |
| Dartmouth Atlas ZipHsaHrr19 | 2019 (latest published) | HSA/HRR |
| ACGME ADS specialty-081 report | AY2026-27 | Program universe |
| SF Match MSDO data report; ACMS match reports | Retrieved 2026-08-18; 2019–2025 | Pipeline |
| CMS Doctors and Clinicians NDF | Modified 2026-07-31 | Graduation years |
| CMS Medicare Monthly Enrollment, annual county and state rows by age group | CY2013–CY2025; June 2026 data release (posted 2026-09-23; retrieved 2026-09-25) | Beneficiaries by age band; fee-for-service shares (S-M10) |
| CMS Medicare Physician & Other Practitioners — by Geography and Service | CY2013–CY2024, national and state rows, HCPCS 17311–17315 (retrieved 2026-09-25) | Trend in cases per beneficiary (S-M10) |
| CMS Medicare Physician & Other Practitioners — by Provider, beneficiary age counts | CY2024 (retrieved 2026-09-25) | Age-specific rates (S-M10) |
| Hauer county population projections by age, sex, and race (SSP2) | Sci Data 2019; OSF project 9ynfc, SSP_asrc.zip (retrieved 2026-09-25) | Aging ratios (S-M10) |
| Weldon Cooper Center national population projections by age and sex | V2026 (July 2026; retrieved 2026-09-25) | Aging sensitivity (S-M10) |

| Source | Vintage / version | Role |
|---|---|---|
| U.S. Census Bureau 2023 National Population Projections, middle series | Released 2023 (retrieved 2026-09-25) | Cross-check of projected 65-and-over totals (S-M10) |

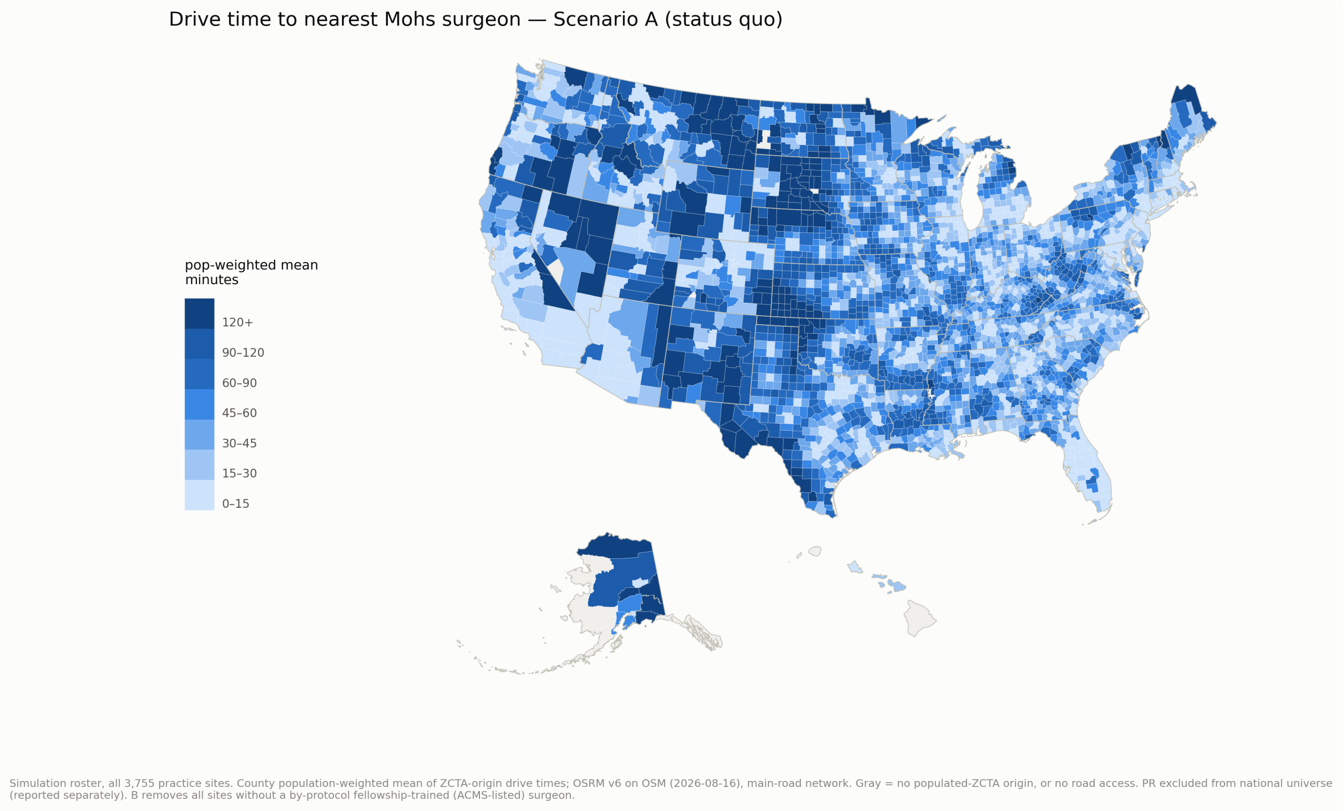


**Figure S1.** Drive time to the nearest Mohs surgeon, status quo (scenario A). Road-network minutes from populated-ZCTA population-weighted centroids; no-road-access origins hatched.

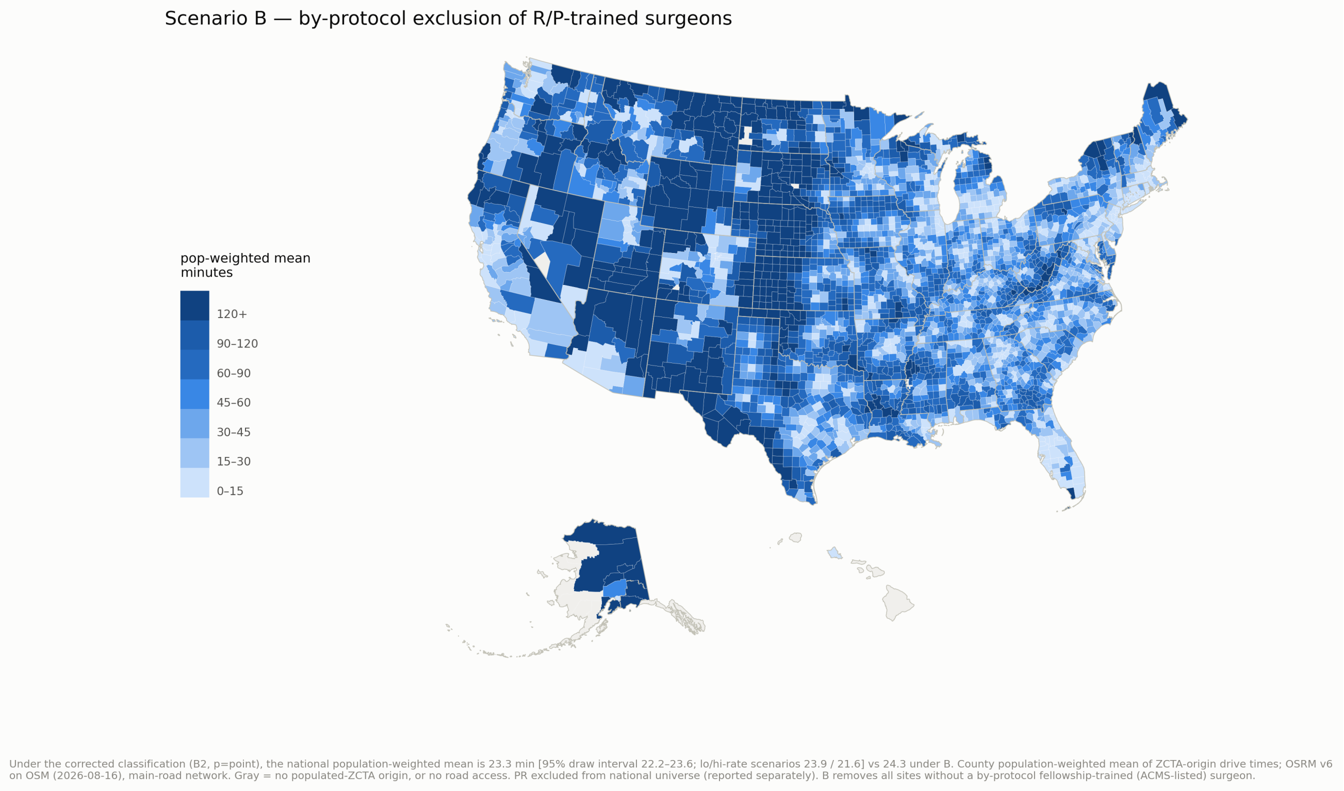


**Figure S2.** Drive time to the nearest Mohs surgeon under by-protocol exclusion (scenario B).

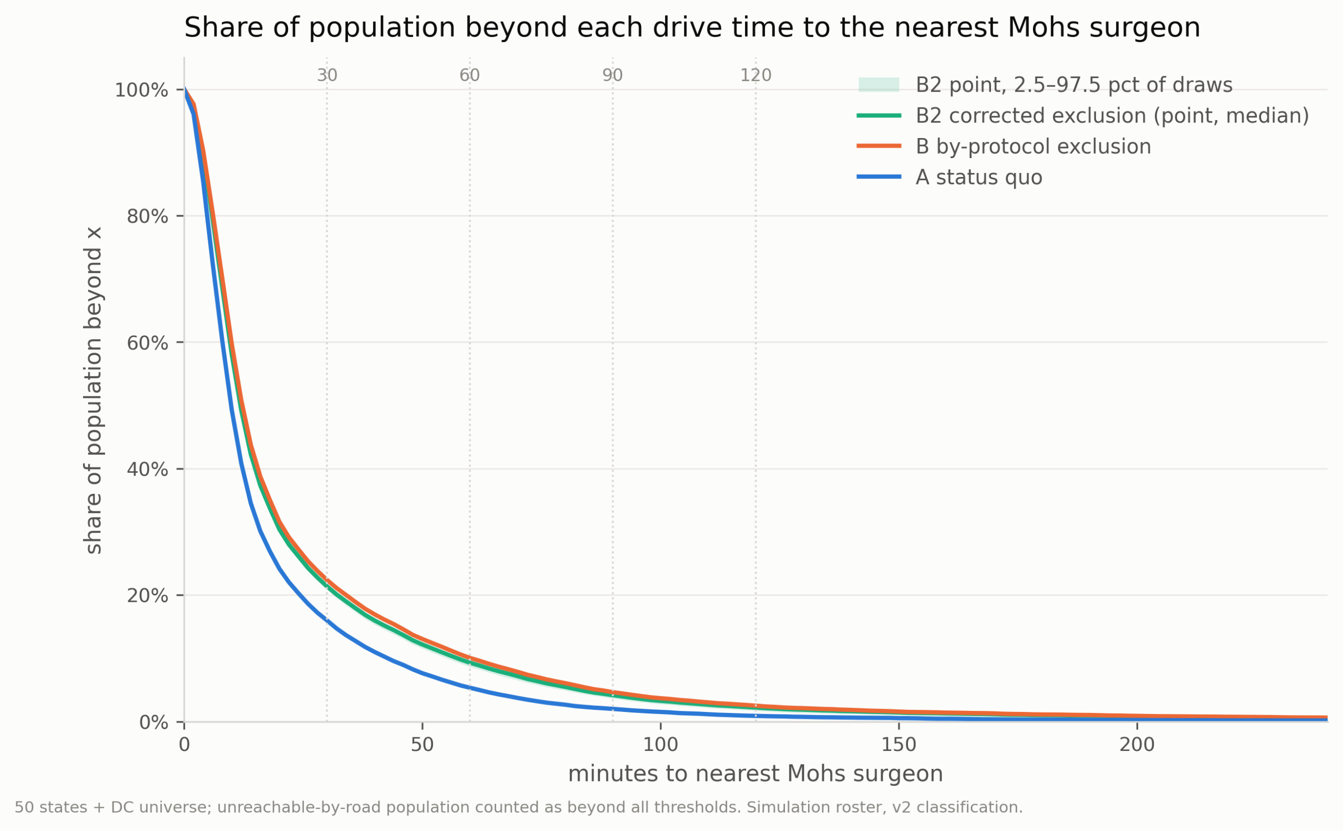


**Figure S3.** Population-weighted survival curves of drive time under scenarios A, B, and B2, with 30/60/90/120-minute thresholds and the no-road-access stratum reported separately.

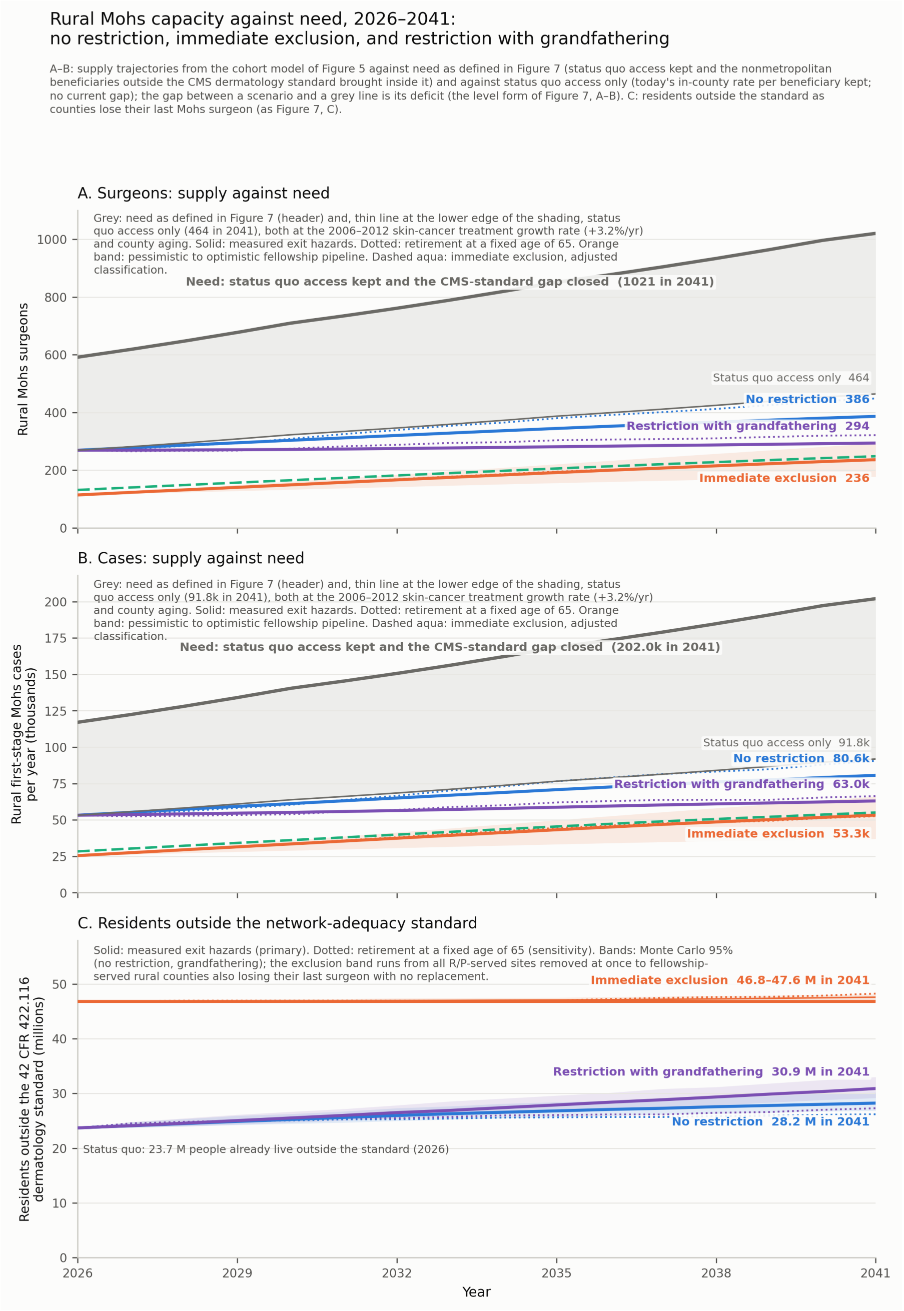


**Figure S4.** Rural Mohs capacity in levels against need, 2026–2041 (the level form of manuscript Figure 7, A–B). A, Rural Mohs surgeons, and B, annual rural first-stage Mohs cases, by scenario (cohort model of Figure 5) against two grey lines, both at the 2006–2012 growth of skin-cancer treatment and county aging. The thick grey line is need as defined in Figure 7, status quo access kept and the nonmetropolitan beneficiaries outside the CMS dermatology standard brought inside it; the thin grey line at the lower edge of the shading is status quo access alone, rural in-county Mohs surgery kept at today's rate per beneficiary with nothing added for residents already outside the standard. Need is 592 surgeons and 117,100 cases in 2026 and 1,021 surgeons and 202,000 cases in 2041 (status quo access alone, 464 and 91,800); in 2041 no restriction reaches 386 surgeons and 80,600 cases per year, a restriction with grandfathering 294 and 63,000, immediate exclusion 236 and 53,300 (adjusted classification 248 and 55,100). C, as manuscript Figure 7, C. Each scenario is drawn twice in its own color, as a solid line in which surgeons exit at the measured hazards (primary analysis) and as a dotted line in which they retire at a fixed age of 65 (sensitivity analysis); the dashed green line is immediate exclusion under the adjusted classification; the band spans pessimistic to optimistic pipeline assumptions in A and B and the Monte Carlo 95% interval in C.

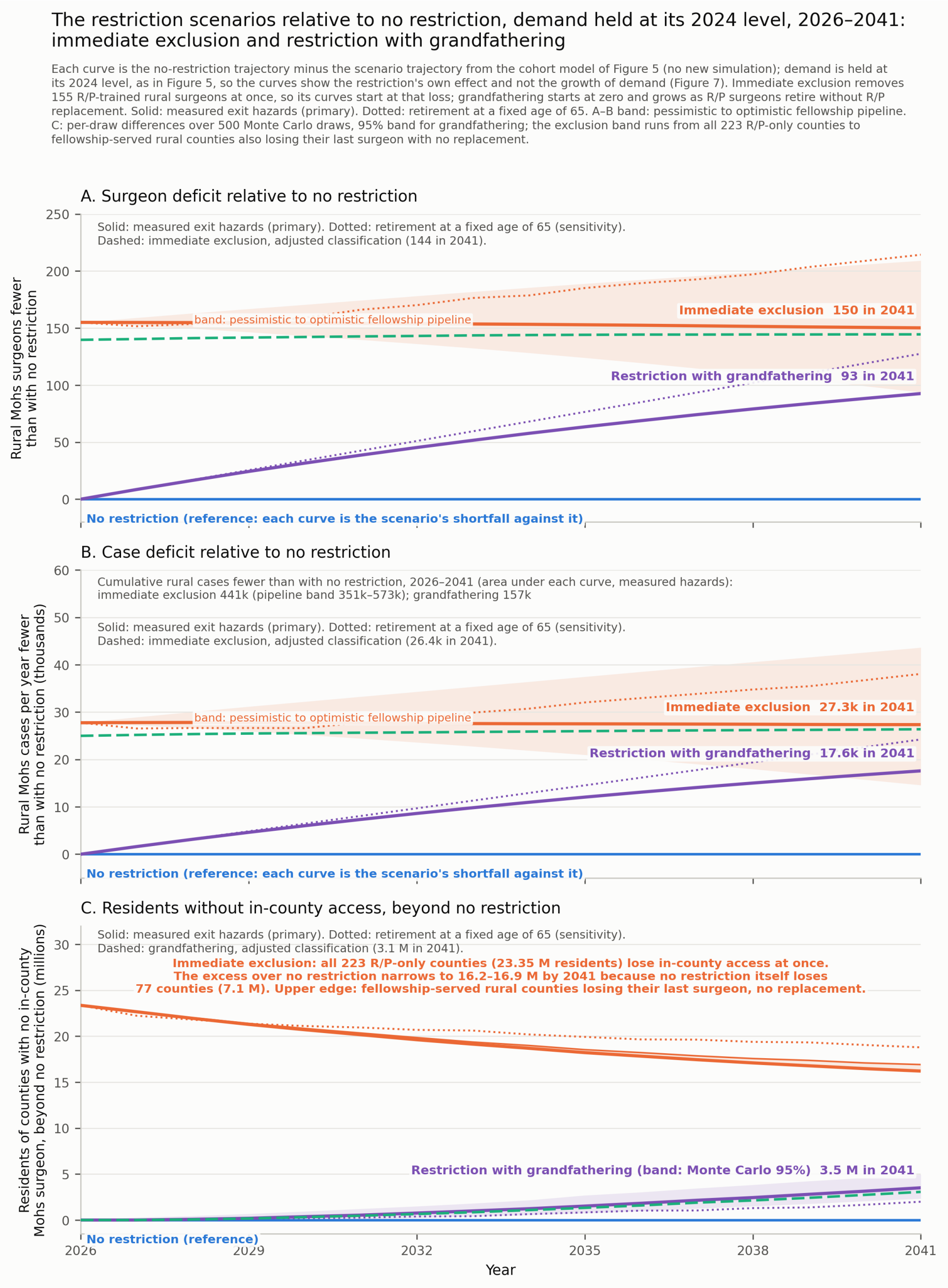


**Figure S5.** The restriction scenarios relative to no restriction with demand held at its 2024 level, 2026–2041 (the difference form of Figure 5). Each curve is the no-restriction trajectory minus the scenario trajectory. A, Rural Mohs surgeons, and B, rural first-stage Mohs cases per year: immediate exclusion starts at the excluded stock (155 surgeons; 27,700 cases) and a restriction with grandfathering grows from zero as R/P surgeons retire unreplaced — 150 surgeons and 27,300 cases per year, and 93 and 17,600, in 2041 (adjusted classification, exclusion: 144 and 26,400); summed over 2026–2041, 441,000 (pipeline band 351,000–573,000) and 157,000 fewer rural cases than with no restriction. C, Residents of counties without an in-county Mohs surgeon beyond no restriction: all 223 R/P-only counties (23.4 million) under immediate exclusion at once, narrowing to 16.2–16.9 million by 2041 because no restriction itself loses 77 counties; a restriction with grandfathering 3.5 [2.3–4.9] million by 2041 (adjusted classification 3.1 million). Each scenario is drawn twice in its own color, as a solid line in which surgeons exit at the measured hazards (primary analysis) and as a dotted line in which they retire at a fixed age of 65 (sensitivity analysis); the dashed green line is the adjusted classification (immediate exclusion in A and B, grandfathering in C); the band spans pessimistic to optimistic pipeline assumptions in A and B and the Monte Carlo 95% interval for grandfathering in C.
